# Executive Functions and ICF Core Sets in Cerebral Palsy: A Systematic Review and Meta-Analysis

**DOI:** 10.64898/2026.02.25.26347013

**Authors:** Alexandra Kalkantzi, Lisa Mailleux, Roser Pueyo, Els Ortibus, Dieter Baeyens, Bernard Dan, Giuseppina Sgandurra, Elegast Monbaliu, Hilde Feys, Saranda Bekteshi

## Abstract

**AIM:** Executive functions (EF) are advanced cognitive processes that play an essential role in daily functioning and may be of increased importance in cerebral palsy (CP), given the complexity of primary and associated impairments. This study aims to synthesize existing evidence on the relation between EF and domains of the International Classification of Functioning, Disability and Health (ICF) in individuals with CP, and to quantify the magnitude of these associations through meta-analysis.

**METHOD:** A systematic literature search was conducted in eight electronic databases up to 14 July 2025, examining associations between EF and ICF domains in CP. EF outcomes were classified into inhibitory control, working memory, cognitive flexibility, higher-order EF, and EF composite scores. Outcome measures were mapped onto ICF domains: Body Functions and Structures, Activity, Participation, and Contextual factors, using the CP Core Sets. Correlation coefficients were transformed to Fisher’s z and entered into three-level meta-analyses to estimate pooled effect sizes. Single moderator analyses examined CP subtype, EF domain, EF assessment type, and mean age. Risk of bias was assessed using the Quality in Prognosis Studies (QUIPS) tool.

**RESULTS:** From 4637 identified records, 38 studies were included, comprising a total sample of 1633 participants with CP. There was substantial heterogeneity in CP subtype, participant age, and EF conceptualization, while the ICF Contextual factors domain was underrepresented. A medium-to-large association was found between EF and functioning across all ICF domains combined (r=0.26, p<0.001). Domain-specific analyses showed a medium association of EF with Body Functions and Structures (r=0.21, p<0.01), a medium-to-large association with Activity (r=0.38, p<0.001) and Participation (r=0.26, p<0.01). CP subtype and mean age significantly moderated the overall EF-functioning association, with mixed CP and younger age associated with stronger effects.

**INTERPRETATION:** EF are meaningfully associated with multiple domains of functioning in individuals with CP. These findings support the relevance of routine EF assessment and suggest that EF are an important cognitive correlate to consider when addressing broader aspects of daily functioning.

**WHAT THIS PAPER ADDS:**

- Executive functions (EF) showed medium-to-large associations with all ICF domains in people with cerebral palsy (CP)
- The strongest and most consistent associations were found between EF and ICF Activity
- Overall associations highlight the relevance of EF as a meaningful intervention target in CP

## INTRODUCTION

With an estimated prevalence of 1.6-3.4/1000 live births and an overall 18 million people worldwide being diagnosed, cerebral palsy (CP) is one of the most common causes of childhood-onset disability.^1^ It comprises a series of non-progressive conditions, attributed to injuries in the developing brain, with direct consequences being movement and posture abnormalities.^2^ Beyond motor impairments, individuals with CP often experience a wide range of non-motor symptoms, such as epilepsy or impairments in somatosensory, cognitive or behavioral function.^3^ The presence and combination of these comorbid signs -together with their varying levels of severity-result in a highly heterogeneous clinical presentation,^4^ characterized by marked interindividual variability in functional abilities. This heterogeneity demands an integrated view of how multiple systems interact to shape everyday performance.

Among the non-motor domains affected in CP, executive functions (EF) play a key role in goal-directed behavior, thereby contributing to overall daily functioning.^5–7^ More specifically, EF comprise a set of advanced cognitive processes that enable individuals to plan, initiate, monitor, and adapt their behavior to achieve purposeful, complex or novel actions.^7^ Various theoretical models conceptualize EF as distinct yet interrelated domains, with the most widely accepted frameworks identifying inhibitory control, cognitive flexibility, and working memory or related aspects^5^ as core components.^7^ Inhibitory control refers to the ability to override prepotent responses and act as appropriate as the situation requires. Cognitive flexibility or shifting involves the ability to adjust to changes, demands and priorities. Lastly, working memory or updating is the ability to retain information that is no longer available (auditory and/or visual) and actively manipulate it in goal-directed behaviour.^7^ Building upon the core components, higher-order EF such as planning, reasoning and problem-solving are developed.^7^ Some other conceptualizations refer to other highly related terms such as self-regulation, and occasionally add concepts to this e.g., attention control.^8,9^ EF are essential not only at the individual level -e.g., academic achievement, adaptive behavior, and individual quality of life-^10,11^ but also at a broader level, by influencing participation and social integration.^12^ Over the past decades, a substantial body of evidence has shown that individuals with CP experience EF difficulties that extend beyond clinical thresholds, with impairments reported across nearly all EF components and across all CP subtypes.^13–16^ Beyond in-depth characterization of EF impairments in CP, research has also shifted towards understanding their relation with other domains of functioning. To date, EF impairments in CP have been associated, among others, with bimanual performance,^14^ psychological functioning,^17^ school functioning, academic performance^18,19^ and quality of life.^11^ Overall, findings demonstrate consistent associations between EF and daily functioning in CP,^11,12,17^ underscoring the relevance of EF in everyday life and highlighting the importance of their early identification and clinical monitoring.

Recent studies have systematically summarized evidence in this field. First, the systematic review and meta-analysis by Zimonyi et al.^20^ quantified EF impairments in CP across studies. Their results showed that people with CP experience significantly more impaired EF across all domains compared to typically developing individuals. Furthermore, Carracedo-Martín et al.^21^ conducted a systematic review regarding the role of cognitive function, including EF, in CP through the lens of the International Classification of Functioning, Disability and Health (ICF).^22^ The ICF conceptualizes health from a biopsychosocial perspective, integrating biological, psychological, and environmental dimensions of functioning. It emphasizes the interaction between Body Functions And Structures, Activities, Participation, and Contextual factors, thereby capturing the complex and multidimensional nature of health and functioning,^22^ even in multi-faceted disorders. In the mentioned systematic review, the relation of cognition was only explored across the ICF domains of Activity, Participation and Environmental factors.^21^ Their results indicated that general intellectual functioning, language and visual perception in CP is associated with multiple ICF chapters, particularly Mobility, Communication and Learning and applying knowledge.

The present review will focus specifically on EF and will explore their relation with all ICF domains, including Body Functions And Structures, Activities, Participation, and Contextual factors. To facilitate the application of the ICF model to specific conditions, ICF Core Sets have been developed to identify the most relevant categories of functioning for particular health populations, including children, teenagers, and adults with CP.^23,24^ The CP-specific ICF Core Sets in this study will serve as a conceptual bridge for examining how EF impairments relate to key components of functioning in CP. Hence, the first aim of this study is to narratively synthesize all available evidence on the relation between EF and ICF sublevels that are included in the CP-specific Core Sets. As a second aim, a meta-analysis will be conducted to estimate the overall effect size (ES) between EF and ICF domains. We hypothesize that worse EF will also result in poorer functioning across all ICF domains. When EF are associated with brain or lesion characteristics, poorer neural integrity is expected to be related with greater executive dysfunction. The last aim is to address the potential clinical and methodological heterogeneity of existing literature through moderator analysis, by examining how specific factors such as EF domain, type of EF assessment tool, mean age, CP subtype play a moderating role in the relation between EF and ICF domains. We hypothesize that CP subtype especially will play a moderating role.

## METHOD

### Protocol development and registration

This systematic review was conducted in accordance with the Preferred Reporting Items for Systematic Reviews and Meta-Analyses (PRISMA) guidelines.^25^ The review was prospectively registered in PROSPERO (CRD42024614057) in November 2024, with minor amendments made in December 2025. Adaptations included: refinement of search strategy; replacement of the PsycINFO database with the APA PsycArticles database; use of Quality in Prognosis Studies (QUIPS) tool for quality assessment of the included studies; use of the R Package for statistical analysis.

### Research question

How are EF related to the ICF Core Sets of Body Functions & Structures, Activity, Participation and Contextual factors in individuals with CP?

### Search strategy

Initial search strategy was conducted by the first author (AK) on 8 February 2025 in eight electronic databases, namely PubMed, EMBASE, MEDLINE Ovid, Cochrane, CINAHL (EBSCO), Web of Science, ERIC IES and APA PsycArticles. The literature search was further updated, resulting in a time frame from inception to 14 July 2025. No search restrictions were imposed for date and language.

The search strategy included keywords focusing on (1) the relevant condition (cerebral pa*, perinatal stroke) AND (2) relation type (relat*, associat*, correlat*) AND (3) study outcomes (executive funct*, OR executive control OR self-regulation OR working memory, inhibition OR inhibitory control, cognitive flexibility). Terms for the ICF were not explicitly defined, to minimize the risk of excluding relevant studies. The Body Functions and Structures domain was considered when EF were associated with movement-related, cognitive, or other fundamental aspects of functioning (e.g., Intellectual function, Mobility of joint functions, Structure of brain). The Activity domain was regarded when EF were associated with aspects assessed through scales, questionnaires, or test batteries that address task execution, mobility, and functional performance (e.g., Acquiring skills, Fine hand use, Walking). Participation was considered in terms of involvement in social contexts, engagement, self-care (e.g., Intimate relationships, School education, Community life), encompassing also the Quality of Life that was examined in relation to psychological state and beliefs. Finally, Contextual factors, included personal (e.g., Social norms, practices and ideologies) and environmental factors (e.g., Health services, systems, policies). Search strategy was tailored to each database using corresponding Boolean operators. The detailed search strategy per database can be found in **Table S1**. Forward search (citation tracking) and backward search (reference list checking) were also implemented in reviews, ensuring comprehensive identification of relevant studies. The retrieved studies were imported into EndNote (Clarivate Analytics, Philadelphia, PA, USA) for deduplication and the remaining records were thereafter uploaded to Rayyan Citation^26^ for screening.

### Eligibility criteria

Inclusion criteria comprised: (1) Participants with a diagnosis of CP. In cases where the study sample consists of a mixed population, at least 80% of participants must have a CP diagnosis to be considered eligible; (2) studies that assessed at least one EF as defined by established frameworks: the models of Miyake and Diamond^5,7^ (i.e., inhibitory control, cognitive flexibility, working memory, problem-solving, reasoning, and planning) or the Anderson model^9^ (i.e., cognitive flexibility, goal setting, attentional control, and information processing); (3) studies that explore the relationship between EF and at least one domain embedded in the CP-specific Core Sets of the ICF; (4) peer-reviewed journal publications including primary studies. No restrictions regarding participants’ chronological age were applied.

### Selection process

Screening was conducted using the blind-on mode of Rayyan.^26^ Title and abstract screening was conducted by the first author (AK) at 100%, while the last author (SB) screened 56% of the records. Across 1.612 overlapping decisioned records, reviewers showed high inter-rater reliability, with 97.6% raw agreement. The full-text screening was then independently repeated at 100% from the first author (AK) and the last author screened 20% of randomly selected records. When characteristics of the study were unclear, study authors were contacted for further clarification. Raw agreement was reached for 91% of the records between the first (AK) and last author (SB). For the remaining 9% of non-agreed records, discussions among the two authors took place, with involvement of a third reviewer (LM) as a consultant, until consensus was reached.

### Data extraction

Data from the included studies was extracted independently by the first author in worksheets. The last author cross-checked the extracted information for all studies and recorded inconsistencies which were resolved through discussion. The following characteristics were extracted from the included studies: Author and publication year, demographic characteristics including sample size, number of females/males, mean age in years and standard deviation, type of CP according to the type of motor disorder (i.e., spastic, dyskinetic, ataxic, mixed) and topographic distribution (e.g., unilateral, bilateral), functional ability based on well-known classification systems, where available (i.e., Gross Motor Function Classification System; GMFCS,^27^ Manual Ability Classification System; MACS,^28^ Communication Function Classification System; CFCS,^29^ Viking Speech Scale: VSS^30^). To ensure consistency when reporting and analyzing the study outcomes, core EF were standardized using Diamond’s and Miyake’s model.^5,7^ Specifically, EF were mapped according to the assessments used in inhibitory control, cognitive flexibility and working memory. Two more categories were coded, namely higher-order EF and EF composite score, reflecting more accurately the EF categorization of the included studies. In accordance with these definitions, the assessment tools used in the included articles were coded, aligning them with specific EF despite the varied terminology used in the original studies. The type of assessment tool of EF (i.e., performance-based test or questionnaire) was also extracted. **Table S2** provides an overview of the tools and the EF domains in which they were categorized.

To classify the ICF domains with which EF have been related, the ICF CP Core Sets were used. The ICF Core Sets for CP are condition-specific selections of ICF categories that capture the most relevant aspects of functioning in this population. They were developed through international, multidisciplinary consensus among clinicians, researchers, and individuals with CP, ensuring that the Core Sets reflect the broad spectrum of challenges and strengths across physical, cognitive, emotional, and social domains.^24^ Distinct Core Sets have been created to represent functioning across the lifespan, including versions for children and youth of different age groups as well as for adults. Two formats are available: a comprehensive version, designed for detailed clinical or research assessment, and a brief version, intended for quicker use in clinical practice or large-scale studies.^23,24^ In this study, the comprehensive versions were used to ensure thorough coverage of all areas of functioning. For each included study, the ICF chapter was reported, as well as categories embedded within the CP-specific Core Set, using the interactive platform *ICF CORE SETS Manual for Clinical Practice* (https://www.icf-core-sets.org/). ^31^ Lastly, correlation coefficients capturing relations between the two aforementioned domains (i.e., EF and ICF Core Sets) were extracted.

### Assessment of study quality

Due to the prognostic nature of the research question, the Quality in Prognosis Studies (QUIPS) tool^32^ was used to evaluate quality of the included studies. This tool has been previously used in systematic reviews and meta-analyses in relevant topics in CP.^20,33^ Risk of bias (RoB) in six domains, including *Study participation, Study attrition, Prognostic factor measurement, Outcome measurement, Study confounding,* and *Statistical analysis and reporting*, was judged qualitatively as low, moderate, or high based on standardized items. For the *Study attrition* domain, although the term does not strictly apply to cross-sectional studies, attrition was considered present when participants were unable to complete part of the assessments, resulting in missing data that were not imputed. Overall RoB was based on qualitative assessment of domain-level judgments. All studies were assessed by the first author (AK) and 20% of the studies from the second author (LM), with a raw agreement of 92%. Any discrepancies were resolved through discussion until consensus was reached in accordance with the predefined criteria of the QUIPS. The template of the QUIPS checklist can be found in **Table S3**.

### Statistical analyses

#### Effect size extraction and preparation

Meta-analysis was conducted to address the magnitude and heterogeneity of the EF-ICF domain association. For this purpose, all reported associations between EF and ICF domains were extracted as correlation coefficients (r) and interpreted descriptively according to Cohen’s d as small (≥0.2), medium (≥0.5), and large (≥0.8).^34^ Due to the heterogeneity of EF and ICF-related assessment tools and their scoring directions, correlation coefficients were sign-adjusted (i.e., reversed when higher scores indicated poorer functioning) prior to analysis to ensure consistent interpretation across studies. All correlation coefficients were transformed into Fisher’s z values. The sampling variance of each ES was calculated as: v_z_ = 1/n−3, where n is the sample size associated with the corresponding correlation. ES were back-transformed to correlation coefficients for interpretation, using the benchmarks proposed by Lipsey and Wilson (<0.10: small; 0.10-0.25: small to medium; 0.25-0.40: medium to large; >0.40 large).^35^

#### Meta-analytic model and handling of effect sizes

As most studies may contribute multiple ES coming within the same sample, ES within studies will be statistically dependent. To appropriately account for this dependency and to avoid underestimation of standard errors, a three-level random-effects meta-analytic model was employed.^36^ This approach decomposes the total variance into three components: Level 1: sampling variance of individual ES, Level 2: within-study variance reflecting dependence among multiple ES derived from the same study, and Level 3: between-study variance reflecting heterogeneity across studies.^36^ Given the inclusion of multiple dependent ES per study and the modest number of studies in several models, cluster-robust variance estimation (CR2) was used to ensure valid statistical inference.

#### Primary meta-analyses and assessment of heterogeneity

First, a grand, three-level random-effects model including pooled ES across all ICF domains was calculated. The Contextual factors domain was excluded from the quantitative analysis due to the small sample of studies (n=3). Thereafter, the same model was run for each ICF domain (i.e., Body Functions and Structures, Activity, Participation). Within each model, all EF-ICF associations corresponding to specific ICF subdomains were included simultaneously. Heterogeneity was quantified using I² statistics. Variance at Level 1 (sampling variance) reflects random error and was summarized using the median sampling variance. Variance at Levels 2 and 3 represent within- and between-study heterogeneity, respectively. I² indicates the proportion of total variance attributable to true heterogeneity (Levels 2 and 3 combined).

#### Sensitivity analyses

Publication bias and small-study effects were examined through Egger’s tests and visual inspection of funnel plots. To account for the potential of publication bias, in significance of the Egger’s test, the trim-and-fill method was used.^37^ This method trims asymmetric studies from the funnel plot, imputes potentially missing studies to restore symmetry, and recalculates an adjusted pooled ES. Further sensitivity analysis was conducted by excluding studies rated as high RoB with the QUIPS^32^ to assess the robustness of the pooled estimates.

#### Moderator analyses

Candidate studies typically use diverse definitions of EF, employ varying neuropsychological measures,^38^ and include heterogeneous samples, making it difficult to compare or generalize results. This methodological variability limits the ability to draw coherent conclusions about the functional consequences of EF impairments in CP. Therefore, moderators such as EF domain, type of tool assessing EF, mean age and CP subtype were used. This analysis was conducted when there was substantial heterogeneity observed in the unconditional models (I²>65%), using three-level mixed-effects meta-analytic models with CR2 inference. The analysis was conducted on the dataset of all ICF domains combined to ensure sufficient statistical power. Moderators were entered individually, and moderation was evaluated either through regression coefficients (for continuous moderators) or through comparisons of subgroup-specific pooled effects using omnibus Wald tests (for categorical moderators). Variables examined included EF domains (i.e., inhibitory control, working memory, cognitive flexibility, higher-order EF, EF composite), EF measurement type (i.e., performance-based task, questionnaire), age (mean age) and CP subtype (i.e., spastic, dyskinetic, mixed, unknown type). Studies that did not specify the CP subtype were excluded from analysis of this moderator. All analyses were conducted in R (RStudio environment) using the *metafor* and *clubSandwich* packages.^39^

## RESULTS

### Search results

The database search and the subsequent stages of the review are reported in a PRISMA flow diagram (**Figure 1**). The search yielded a total of 4637 references, 1157 of which were duplicates. The forward and backward citation tracking of relevant reviews did not yield additional studies that fit with the present review’s scope. After deduplication, 3480 studies were screened based on titles and abstract, out of which 3391 were excluded. Eighty-nine studies were sought for full-text retrieval, 13 of which were excluded due to being duplicate (n=2), conference proceedings (n=9), and no full text available (n=2). Seventy-six studies were retained for full-text screening, out of which 38 were excluded with reasons for exclusion reported in **Table S4**. Finally, 38 studies^14,16–18,40–73^ were included in this review and can be found in **Table S5**.

**Figure 1.**
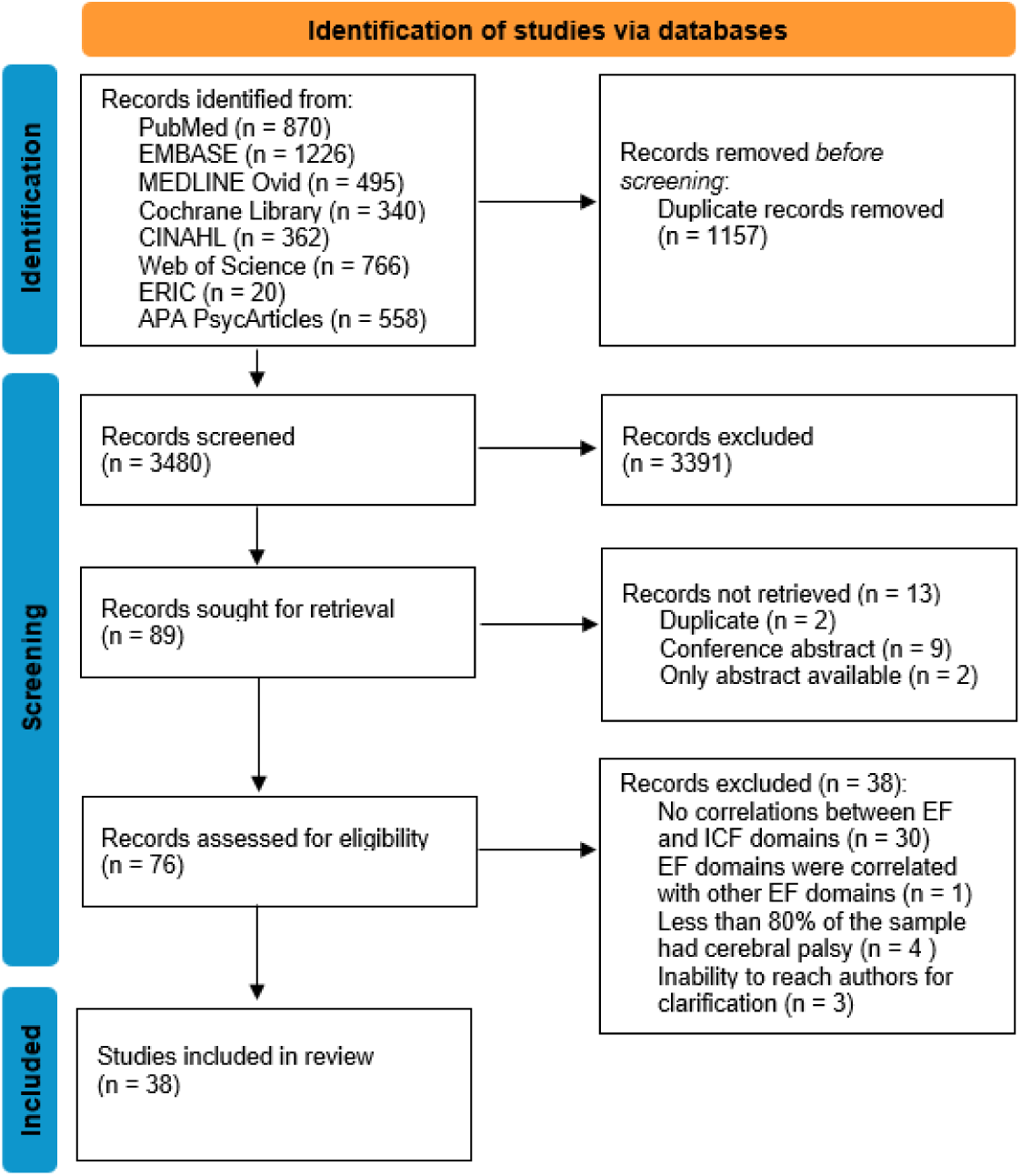
PRISMA 2020 flow diagram for searches of databases Abbreviations: EMBASE, Excerpta Medica dataBASE; CINAHL, Cumulative Index to Nursing and Allied Health Literature; ERIC IES, Education Resources Information Center Institute of Education Sciences; EF, executive functions; ICF, International Classification of Functioning, Disability and Health. Source: Page MJ, et al. BMJ 2021;372:n71.

### Participant characteristics

Across the included studies, a total of 1633 participants were included, and among the 1556 (95.3%) participants with available sex data, 665 (42.7%) were female and 891 (57.3%) were male. The included age ranged from 3 to 62 years, based on all studies reporting age information. In terms of mean age, participants in three studies^48,65,72^ were aged 0-6 years, 28 studies^14,16–18,40,42,44–47,49,51–54,58–64,66–71^ included participants aged 7-12 years, two studies^43,73^ involved adolescents aged 13-18 years, and five studies^41,50,55–57^ included adults older than 18 years. Motor type of disorder was reported for 90.3% of the total sample, that is, for 1474 participants. Across all studies, spastic CP was the most common presentation, followed by dyskinetic types. To obtain an overall pooled estimation of CP subtype distribution, individual subtype counts were summed across all studies, resulting in the following pooled distribution: 1210 participants with spastic CP, 216 with dyskinetic CP, 24 with ataxic CP, 20 with mixed motor types and 4 with unknown CP type. Motor type was not reported for the remaining 159 participants. When classifying study-level samples based on the type of motor disorder, 15 studies (39.5%) ^14,16–18,40,43,46–48,50,58,61,62,64,66^ included only participants with spastic CP, four studies (10.5%) included^41,55–57^ only dyskinetic CP, 14 studies^44,45,49,51–53,59,63,65,67,69–72^ included mixed motor types (e.g., people with spastic, dyskinetic and ataxic type; 36.8%) and in 5 studies (13.2%) ^42,54,60,68,73^ the type of motor disorder was not explicitly mentioned. No studies included pure ataxic types. Functional classification was available for only a subset of the total sample, that is, reported in 21/38 studies for a total of 964 participants (59% of the total sample). Specifically, 19/21 studies reported GMFCS, 15/21 studies reported MACS, 7/21 studies mapped CFCS, 3/21 studies reported VSS, and 2/21 studies reported BMFM levels. In the text, distributions are reported by synthesizing classification data into two severity groups: levels I-III, reflecting ambulatory or lower-severity functioning, and levels IV-V, reflecting non-ambulatory or higher-severity functioning. An exception was made for the VSS, which is reported according to its original four-level structure. The GMFCS was the most frequently reported, with data available for 856 participants. Of these, 634 individuals (74.1%) were classified in GMFCS levels I-III, while 222 (25.9%) were classified in levels IV-V. The MACS was reported for 619 participants, with the majority classified in MACS levels I-III (n=548; 88.5%), and a smaller proportion in levels IV-V (n=71; 11.5%). The CFCS data were available for 302 participants, of whom 268 (88.7%) were classified in CFCS levels I-III and 34 (11.3%) in levels IV-V. The VSS was reported for 94 participants, most classified in VSS levels I-II (n=64; 68.1%), while 30 participants (31.9%) were classified in levels III-IV. The BMFM was available for 112 participants, with 94 individuals (83.9%) classified in levels I-III and 18 (16.1%) in levels IV-V. Functional classification data were unavailable or unreported for 16 participants across the 21 studies. Reporting of age, functional classifications, and motor type was inconsistent across studies, and the summary of the above demographic characteristics only reflects data from studies where the relevant information was available. Finally, several studies reported co-occurring conditions commonly associated with EF impairments, including attention-deficit/hyperactivity disorder (ADHD; n=6)^14,17,47,54,58,62^ and autism spectrum disorder (ASD; n=4).^14,17,49,54^ Additionally, two studies^67,68^ excluded participants with ASD diagnosis.

### Descriptive characteristics of the included studies

Of the included studies, 29^14,16–18,40,42–44,46–48,50,54–64,66,68–70,72,73^ had a cross-sectional design_65,67_ Furthermore, included were six longitudinal studies,^51–53,65,67,71^ one case-control study,^41^ one group case study,^45^ and one randomized controlled trial.^49^ For the last study, only the baseline measures were considered. EF domains were assessed in various combinations, with studies assessing only one domain of core EF, two or all core EF, core and higher-order EF, only higher-order EF, only EF composite score or combination of core, higher-order and composite EF. More specifically, 15 studies (39.4%)^14,16,18,42–44,48,49,52,53,57–60,66^ included measures of inhibitory control, 18 studies (47.4%)^14,18,44–46,48–53,58,59,63,65,70–72^ included measures of working memory, 15 studies (39.4%)^14,16,18,41,48,49,52,53,55–60,66,70,71^ included measures of cognitive flexibility, 10 studies (26.3%)^14,18,46,49,56–58,66,68,73^ assessed higher-order EF and 13 studies (34.2%)^14,17,18,40,47,54,58,61,62,64,67,69,73^ reported an EF composite score. Regarding the type of tools used to assess EF, 25 studies (65.7%)^16,41,43–46,48–53,55–57,59,60,63,65,66,68–72^ used only performance-based tools, 12 studies (31.6%)^14,17,18,40,42,47,54,58,61,62,64,67^ used only a questionnaire, and one study (2.7%)^73^ used both performance-based tools and a questionnaire.

#### EF-ICF Body Functions and Structures

The frequency of ICF chapters in the included studies are presented in **Figure 2**. Twenty-two _studies_^16,17,42–44,46,47,50,55–59,61,62,64,65,67–69,71,72^ explored associations between EF and the ICF Body Functions and Structures domain (**Table 1**). Of those, 15 studies ^17,42–44,47,50,56,59,61,65,67–69,71,72^ reported associations within the ICF chapter Mental functions (b1). This chapter involves brain functions related to consciousness, orientation, perception, emotion, thought and behaviour. Outcome measures of two studies^50,65^ were also embedded under the ICF chapters Sensory functions-pain (b2) and two^65,67^ under the Voice and speech functions chapter(b3). Chapter b2 refers to functions related to sensing stimuli and experiencing pain, whereas b3 encompasses functions such as voice quality, pitch and fluency. Six studies ^16,55,57,58,62,64^ explored brain lesion characteristics, embedded in the ICF chapter Structures of the nervous system (s1). Lastly, one study explored dental caries, embedded under the chapter Structures involved in voice and speech (s3).^46^

#### EF-ICF Activity

Fourteen studies^14,41,45,47,51–53,60,66,67,70–73^ explored associations between EF and the ICF Activity domain (**Table 1**). Nine of these studies ^45,47,51–53,60,70–72^ included measures within the ICF chapter Learning and applying knowledge (d1), related to learning, thinking and applying acquired knowledge. Communication (d3) at the level of activity was assessed in one study^41^ through the CFCS. Lastly, eight studies ^14,47,66,67,70–73^ assessed modalities embedded within the Mobility chapter (d4), which encompasses activities such as moving and changing body position or location (i.e., walking, transferring, using transportation, hand-arm use).

**Figure 2.**
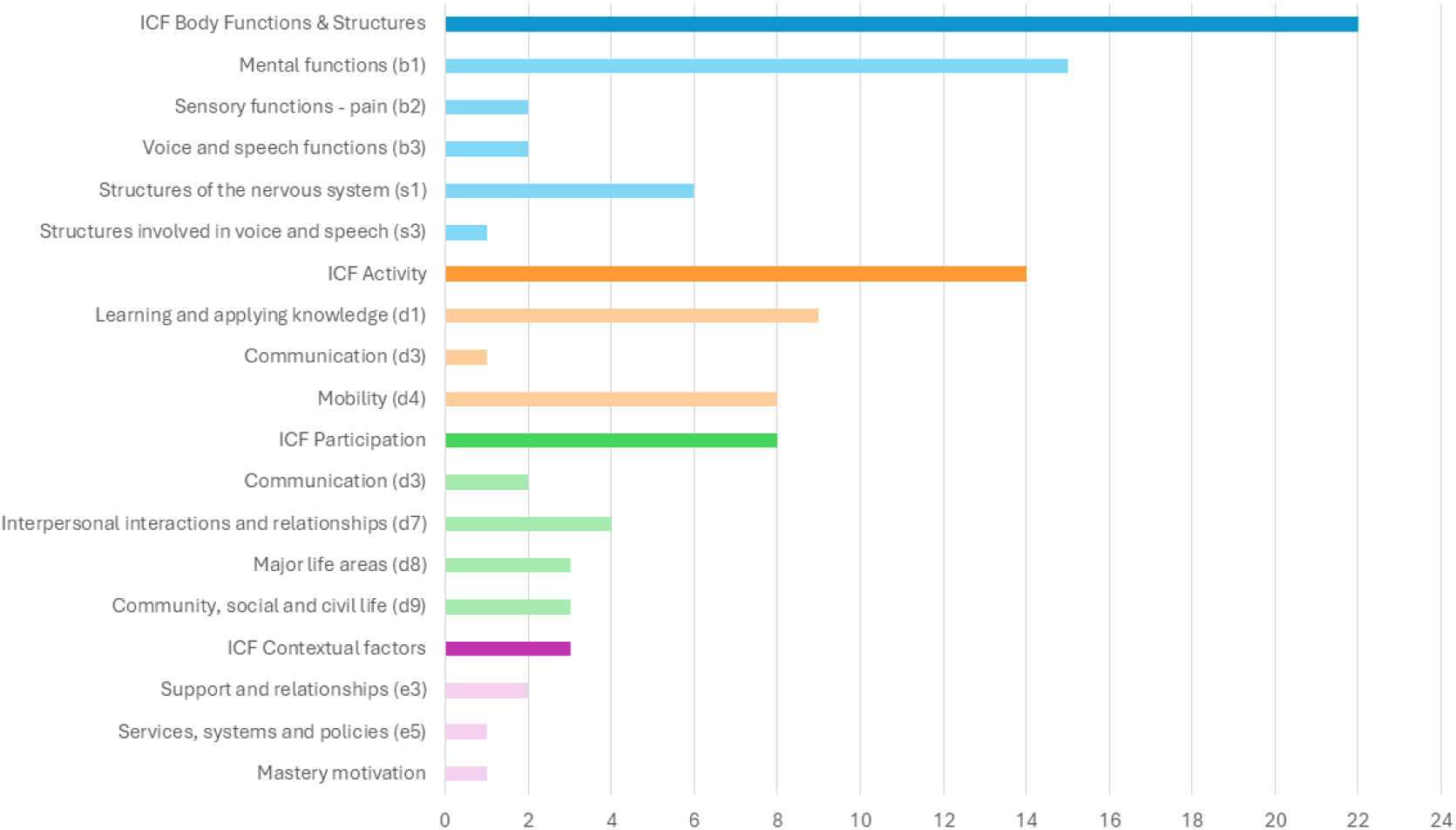
Frequency of ICF chapters examined in the included studies

#### EF-ICF Participation

Eight studies^18,40,47–49,56,63,73^ reported associations between EF and ICF Participation domain (**Table 1**). The ICF chapter Communication (d3) was assessed in two studies^56,63^ involving activities related to understanding and producing messages through spoken, written, signed, or alternative communication. Four studies^40,47,49,73^ assessed Interpersonal interactions and relationships (d7), reflecting interactions with family, friends, colleagues, and intimate relationships. The chapter Major life areas (d8), which includes activities related to education, work, employment, and economic life was assessed in three studies.^18,40,56^ Community, social and civil life (d9), encompassing activities related to participation in community life, recreation, leisure, religion, and civic or political life was assessed in three studies.^40,48,56^

#### EF-ICF Contextual factors

Lastly, three studies^54,56,73^ explored the relation between EF and ICF Contextual factors (Personal and Environmental). Two of these studies used measures embedded in the ICF chapter Support and relationships (e3),^54,56^ with one of them also addressing the ICF Services, systems and policies (e5) chapter.^56^ The third study explored mastery motivation,^73^ which consists of a personal factor and does not obey to a specific ICF chapter. Important to note that each study may have explored more than one ICF chapters.

### Synthesis of results

#### Associations between executive functions and ICF Body Functions and Structures

A thorough overview of the associations in each study, classified per ICF domain and chapter can be found in **Table 1**. Significant associations with *inhibitory control* were primarily found in ICF Mental and perceptual function chapter (b1). Li et al.^59^ reported that poorer *inhibitory control* was correlated with worse social-cognitive function; specifically, higher error rates on *inhibitory control* were associated with lower theory-of-mind scores on false-belief (r=-0.58) and faux pas (r=-0.37). Cabezas & Carriedo^43^ found that errors on a similar task were negatively correlated with temporal processing speed generalization (r=-0.60). Brain lesion characteristics, embedded within the ICF chapter Structures of the nervous system (s1) also showed a significant relation with *inhibitory control*. Larsen et al.^58^ found that higher parent-rated *inhibitory control* problems correlated with increased axial diffusivity in frontal white matter tracts (rs=0.58), suggesting that poorer *inhibitory control* was associated to microstructural abnormalities. Laporta-Hoyos et al. 2018^57^ further found small correlations between *inhibitory control* and parietal lobe score (r=0.24) and medial dorsal thalamus (r=-0.29).

Furthermore, significant associations were consistently observed between *working memory* and the ICF Mental function chapter (b1), particularly general intellectual functioning and language. Van Rooijen et al. 2015^71^ found that digit span backward correlated positively with nonverbal reasoning (r=0.61) and language comprehension. Similarly, another study by van Rooijen et al. 2016^72^ reported that backward digit recall was significantly associated with both nonverbal IQ (r=0.52) and language proficiency (r=0.48). Using a memory updating paradigm, Li et al.^59^ found moderate-to-high positive correlations with theory-of-mind (false-belief r=0.73; faux pas recognition r=0.64). Peeters et al.^65^ likewise noted that *working memory* was positively related to general intelligence (r=0.48) and phonological processing (first-phoneme recognition, auditory discrimination, word articulation; r=0.57, r=0.63, r=0.73, respectively), embedded in the ICF chapters Mental functions (b1), Sensory functions and pain (b2) and Voice and Speech functions (b3). Structures of the nervous system (ICF chapter s1) also showed multiple significant correlations with *working memory*. Larsen et al.^58^ found that greater parent-rated *working memory* impairments were associated with higher diffusion in white matter tracts (mean diffusivity r_s_=0.51, axial diffusivity r_s_=0.50). On the ICF chapter Structures involved in voice and speech (s3), La Rocha Dourado et al. found that lower scores in the digit span backward was associated with more teeth cavities (r=-0.42).^46^

Fewer robust correlations were noted for *cognitive flexibility* measures. In the Mental function’s ICF chapter (b1), Laporta Hoyos et al. 2017^56^ reported that better *cognitive flexibility* was associated with better feelings about functioning (r_s_=0.51). In studies related with the chapter Structure of the nervous system (s1), both Di Lieto et al.^16^ and Laporta Hoyos et al. 2018^57^ reported significant associations between *cognitive flexibility* and corpus callosum (r=-0.55 and r=0.39, respectively). Laporta Hoyos et al. 2018,^57^ further reported worse *cognitive flexibility* associated to frontal (r=0.34), parietal (r=0.25), and temporal (r=0.25) lobe, as well as reduced integrity of the posterior thalamus (r=-0.44).

*Higher-order EF* also showed significant associations with the relative ICF domain. Stadskleiv et al.^68^ found that planning correlated with language skills; specifically, with expressive verbal ability (r=0.60) and language comprehension (r=0.40). Structural brain correlates (ICF chapter s1) were also evident for *higher-order* tasks. Laporta-Hoyos et al. 2018^57^ found that planning was inversely related to overall brain lesion volume (r=-0.32), frontal lobe lesion extent (r=-0.32), parietal (r=-0.24), temporal (r=-0.34), occipital lobe (r=-0.29), posterior thalamus (r=-0.25), caudate (r=-0.32) and corpus callosum (r=-0.41).

Regarding relations with the *EF composite*, examining the chapter Mental functions (b1) and specifically the subdomains perception, emotional, thought and psychomotor functions, Whittingham et al.^17^ found that overall executive dysfunction was associated with more conduct (r=-0.43) and hyperactivity problems (r=-0.49), and with fewer prosocial behaviours (r=0.28). Similarly, Li et al.^61^ found that *EF composite* was positively related to emotional and thought functions (r=0.67). Further, Forsman & Eliasson^47^ reported moderate to high correlations of the *EF composite* with Mental functions (b1) of emotions, language, memory and perception (r values ranged between 0.42 to 0.81).

#### Associations between executive functions and ICF Activity domains

Few significant correlations were found between inhibitory control and the ICF Activity domain. Within the chapter Learning and applying knowledge (d1), Li et al. found that *inhibitory control* was significantly correlated with calculation (r=0.57) and applied problems (r=0.75). In the ICF Mobility chapter (d4), Kalkantzi et al.^14^ found that better BRIEF *Inhibition* scores were correlated with better bimanual performance (r values ranged between -0.34 and -0.48).

*Working memory* was repeatedly linked to competence in school-related activities, especially numeracy, embedded within the ICF chapter Learning and applying knowledge (d1). Jenks et al. 2007^51^ found that digit span backward correlated with basic math skills, such as counting (r=0.63), simple addition (r=0.65), simple subtraction (r=0.54), and conceptual number knowledge (r=0.50). Critten et al.^45^ also reported that backward digits correlated with reading ability (r = 0.63) and nonsense passage reading (r=0.64). Van Rooijen et al. 2015^71^ found that backward digit span correlated positively with arithmetic performance (ICF chapter Learning and applying knowledge; d1) and motor activities (ICF chapter Mobility; d4), that is, backward span with arithmetic (r=0.37), gross motor (r=0.40) and fine motor skills (r=0.41). In a follow-up by the same group,^72^ digit recall backward correlated with early numeracy at 6 (r=0.63) and 7 years (r=0.66). Other significant associations of *working memory* are reported in **Table 1**.

At the level of Activity, *cognitive flexibility* was also mainly associated with academic skills (Learning and applying knowledge; d1). Specifically, *cognitive flexibility* was correlated negatively with reading performance (r=-0.35),^53^ and math, including simple subtraction reaction time (r=0.32) and complex addition reaction time (r=0.41),^52^ based on the studies of Jenks et al. *Higher-order EF* were mainly related to functions at the ICF Mobility chapter (d4). Warschausky et al.^73^ noted a negative correlation between a planning task and manual ability (r=-0.38), but no other significant correlations. Additionally, poorer planning and organization was correlated with poorer bimanual performance (r ranged between -0.31 and -0.35).^14^

Lastly, *composite EF* measures were also mainly related to ICF Mobility chapter (d4). Forsman and Eliasson reported associations between *EF composite* and general learning and motor activity, with specific correlations corresponding to adaptive learning (r=0.76), gross motor (r=0.43) and fine motor skills (r=0.70).^47^ Kalkantzi et al.^14^ found close-to-moderate associations between parent-rated BRIEF total and bimanual performance (r values ranged between -0.42 and -0.48). Two studies found no significant associations between *EF composite*, GMFCS and MACS.^67,73^

#### Associations between executive functions and ICF Participation domains

Several studies found that core EF (i.e., inhibitory control, working memory, cognitive flexibility) were significantly related to chapters of ICF Participation. *Inhibitory control* was mainly related to Major life areas (d8), particularly school participation (r ranged between 0.19 to 0.31).^18^ Nordberg et al.^63^ found no correlations between *working memory* and Communication (ICF chapter d3), specifically grammar comprehension, vocabulary, and a storytelling measure. Nevertheless, in the study of Mousavi et al.^18^ *working memory* showed small correlations with school participation (r=0.22), manipulation with movement (r=0.19) and setup/cleanup (r=0.21). *Cognitive flexibility* also showed small associations with overall school participation (e.g., participation, travel/movement tasks), with coefficients ranging from r=0.18 to 0.39.^18^ *Cognitive flexibility* further showed a moderate positive correlation with Communication (ICF chapter d3); particularly, preservative errors were associated with general communication and physical health (r=0.40).^56^ In the same study of Laporta-Hoyos et al. 2017,^56^ *cognitive flexibility* correlated also with Major life areas (ICF chapter d8), specifically general well-being (r=0.36). Mousavi et al.^18^ found associations between *cognitive flexibility* and school participation (ICF chapter Major life areas; d8), with the coefficients ranging between r=0.18-0.39.

*Higher-order EF* were mainly related to ICF chapters Major life areas (d8) and Community, social and civic life (d9). Laporta-Hoyos et al.^56^ found that both core and *higher-order EF* measures correlated with CP-specific quality-of-life domains. Using the Tower of London task, Warschausky et al.^73^ found that planning was correlated with daily-living skills (r=0.43). Further, Mousavi et al.^18^ provided a detailed profile of EF-school participation associations, namely planning correlated with overall school participation (r=0.39) and with classroom setup tasks (r=0.35). BRIEF Planning and Organization correlated with hygiene (r=0.47) and clothing management (r=0.31) Looking at relations between *EF composite* scores and ICF Participation, Akyurek et al.^40^ reported that EF-routines correlated with outcomes embedded in the ICF chapters Interpersonal interactions (d7),

Major life areas (d8) and Community, social and civic life (d9). Specifically, *EF composite* scores were related to self-regulation skills (r=0.63), communication skills (r=0.62) and school-related functions (r=0.71). Warschausky et al.^73^ reported that *EF composite* correlated with adaptive social and conceptual skills (Interpersonal interactions and relationships; d7), that is, with conceptual adaptive composite (r=0.56) and social composite (r=0.61). In the same ICF chapter, Forsman & Eliasson^47^ found that the *EF composite* associated significantly with social competence (adaptive behavior; r=0.64). García-Galant et al.^49^ found that both core EF composite and higher-order EF correlated with social cognition (Interpersonal interactions and relationships; d7; r=0.47 and r=0.37, respectively). Lastly, Mousavi et al.^18^ reported links between *EF composite* and multiple subscales, such as participation (r=0.40) and hygiene (r=0.48), embedded in the ICF chapter Major life areas, d8.

#### Associations between executive functions and ICF Contextual factors

Two studies explored associations between EF and ICF contextual factors, namely Support and relationships (ICF chapter e3; **Table 1**). Laporta-Hoyos et al. 2017^56^ reported a small correlation between *cognitive flexibility* and family health (rs=0.31). Additionally, the study of Warschausky et al. found another small correlation between *EF composite* and mastery motivation (r=-0.44). No other significant correlations were reported in this ICF level.

**Table 1.**
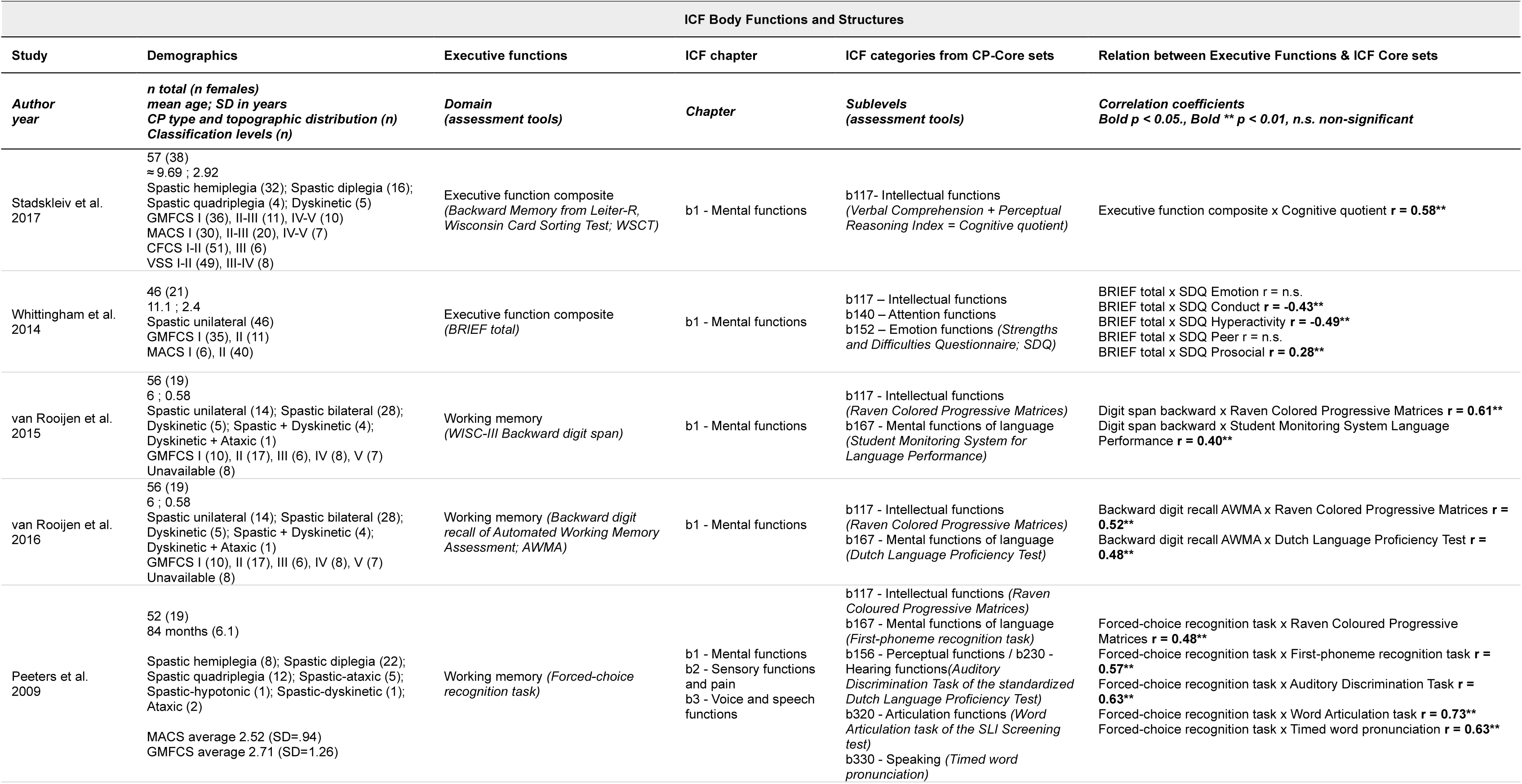
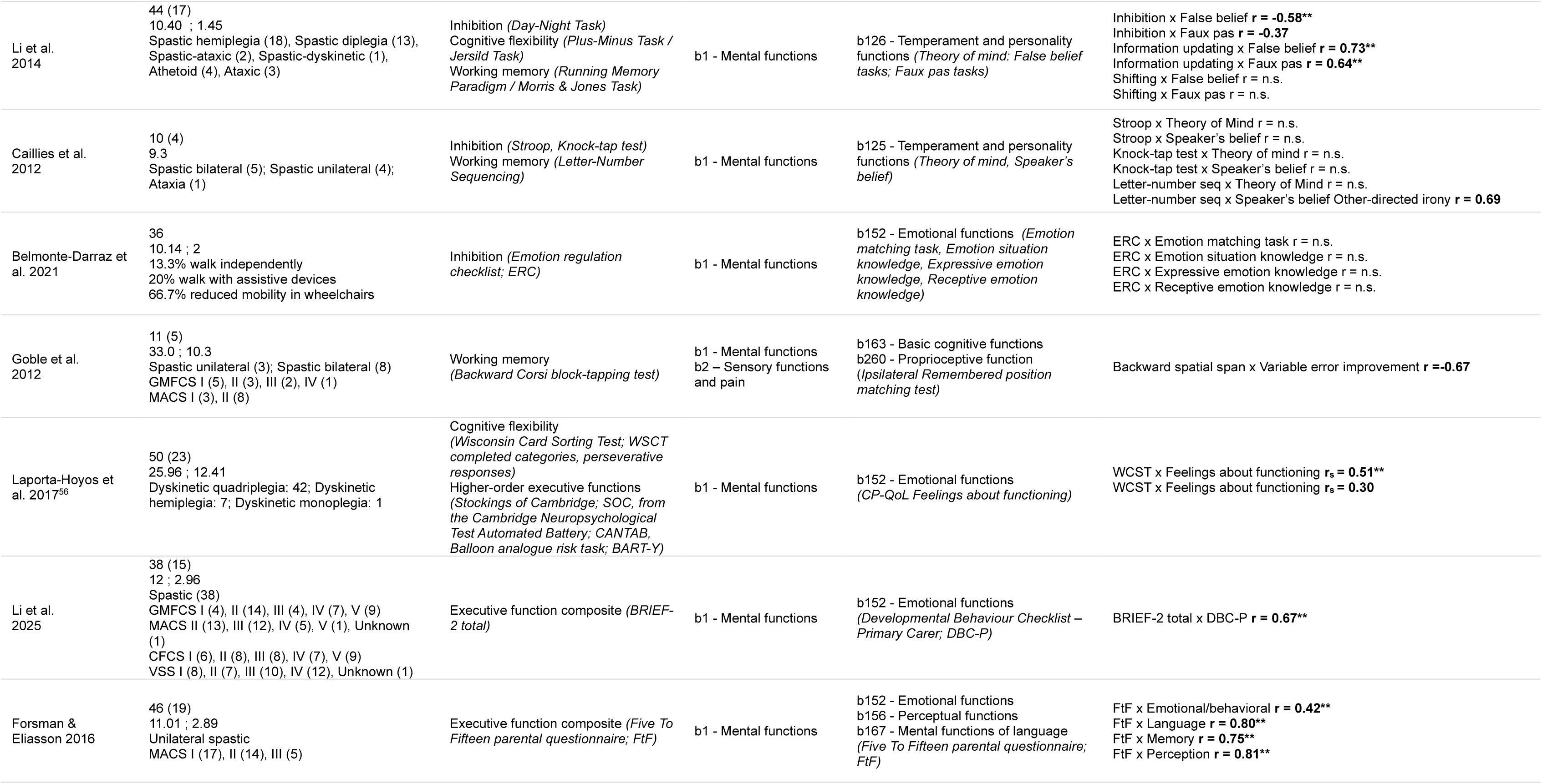
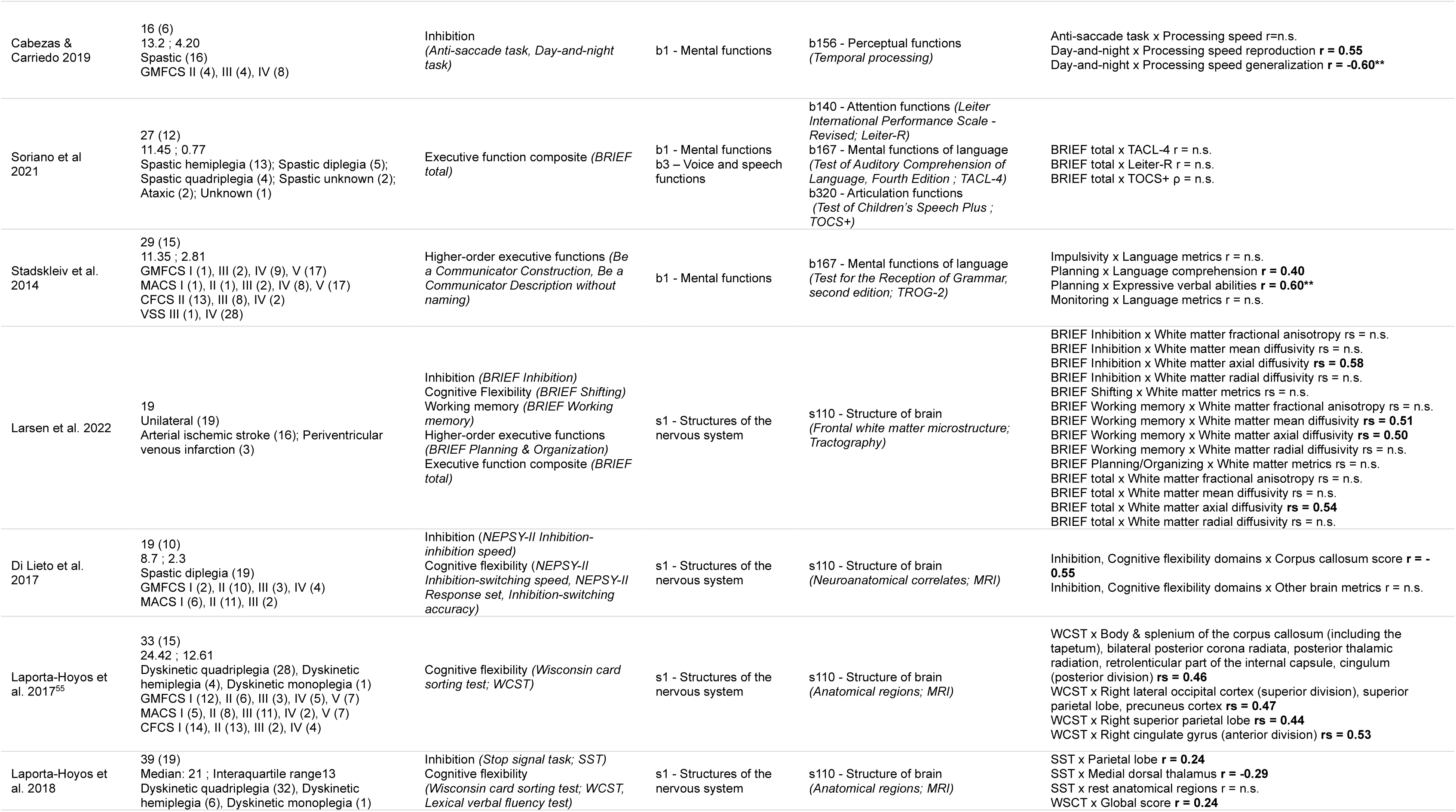
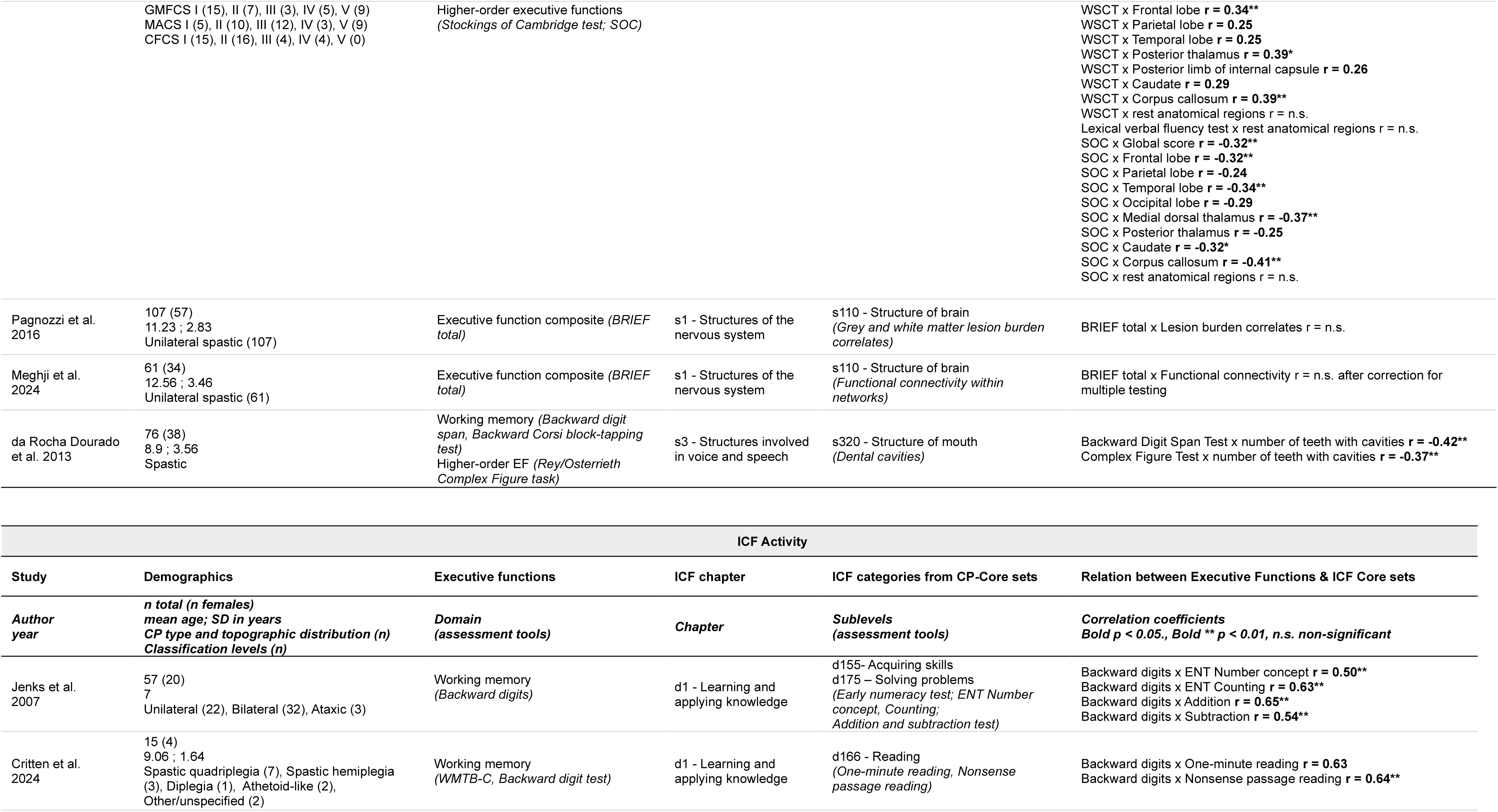
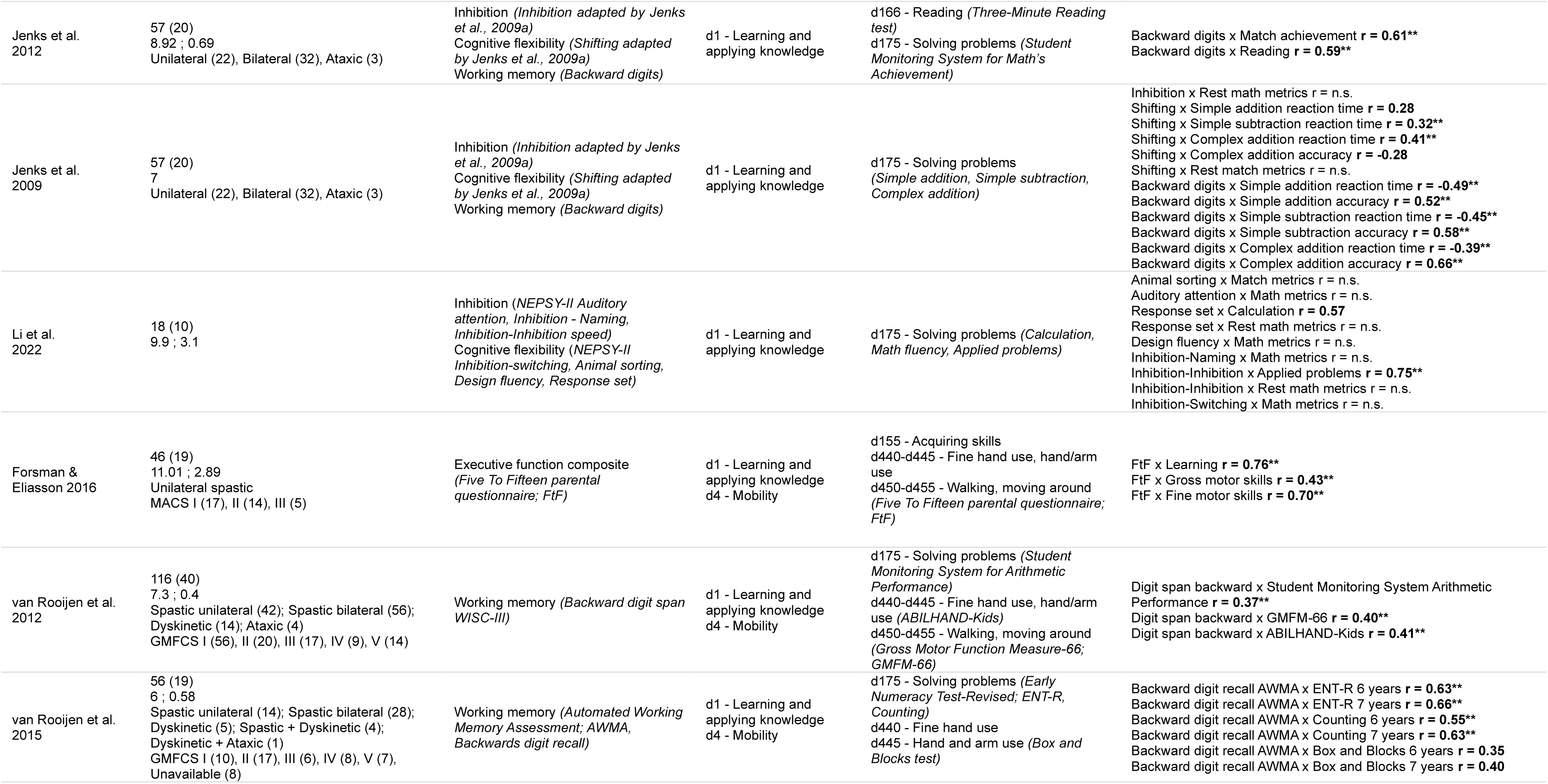
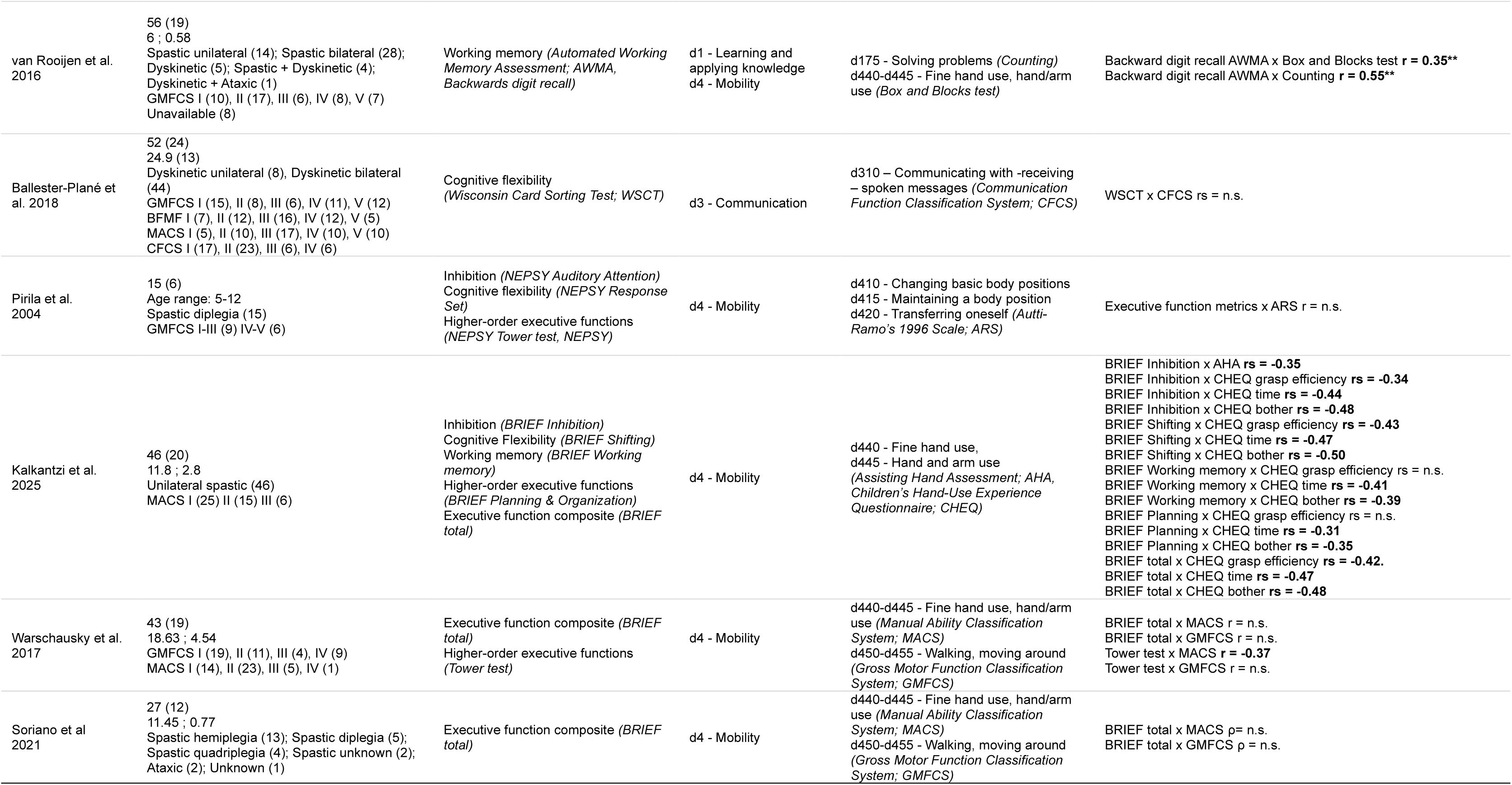
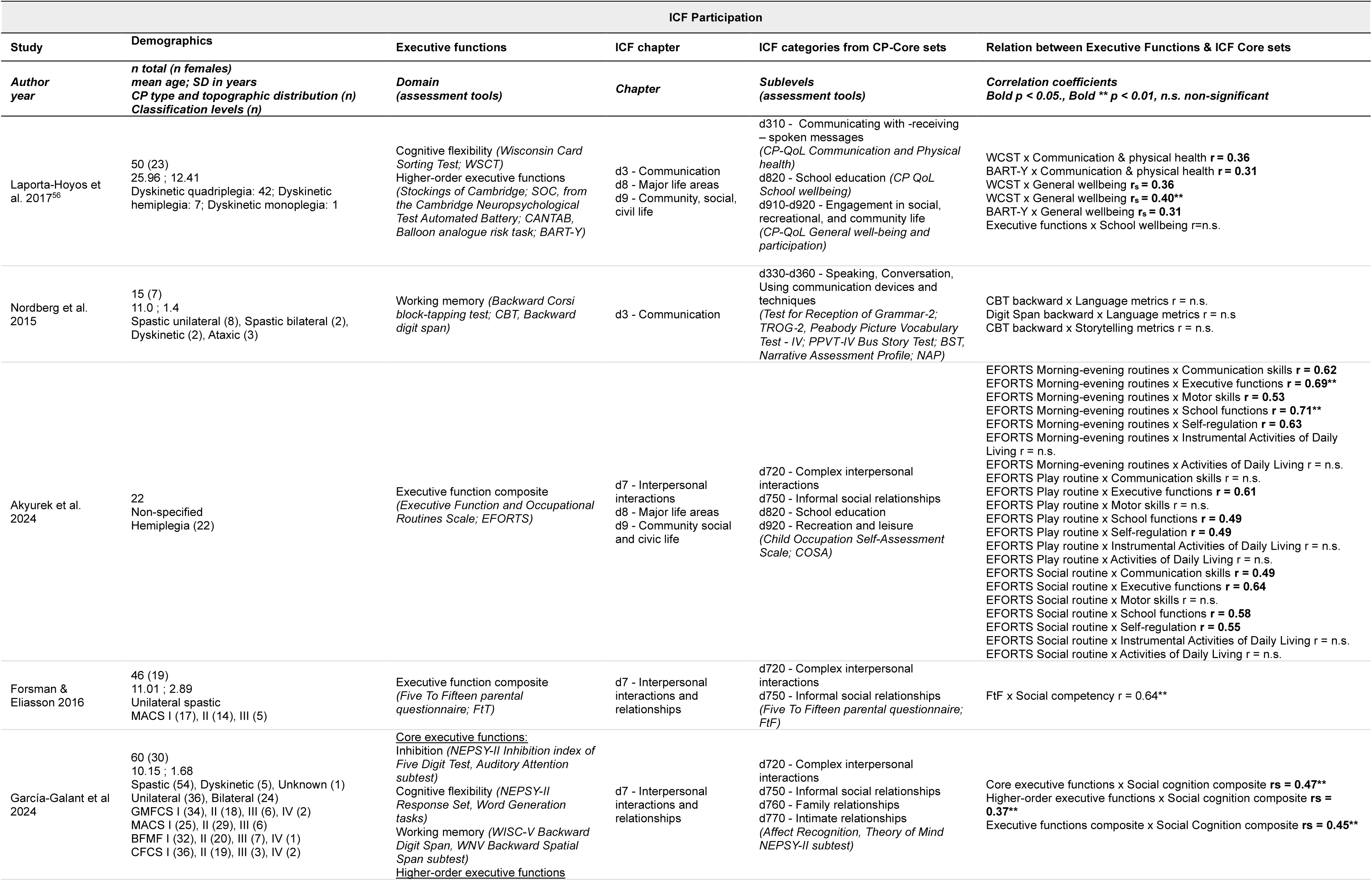
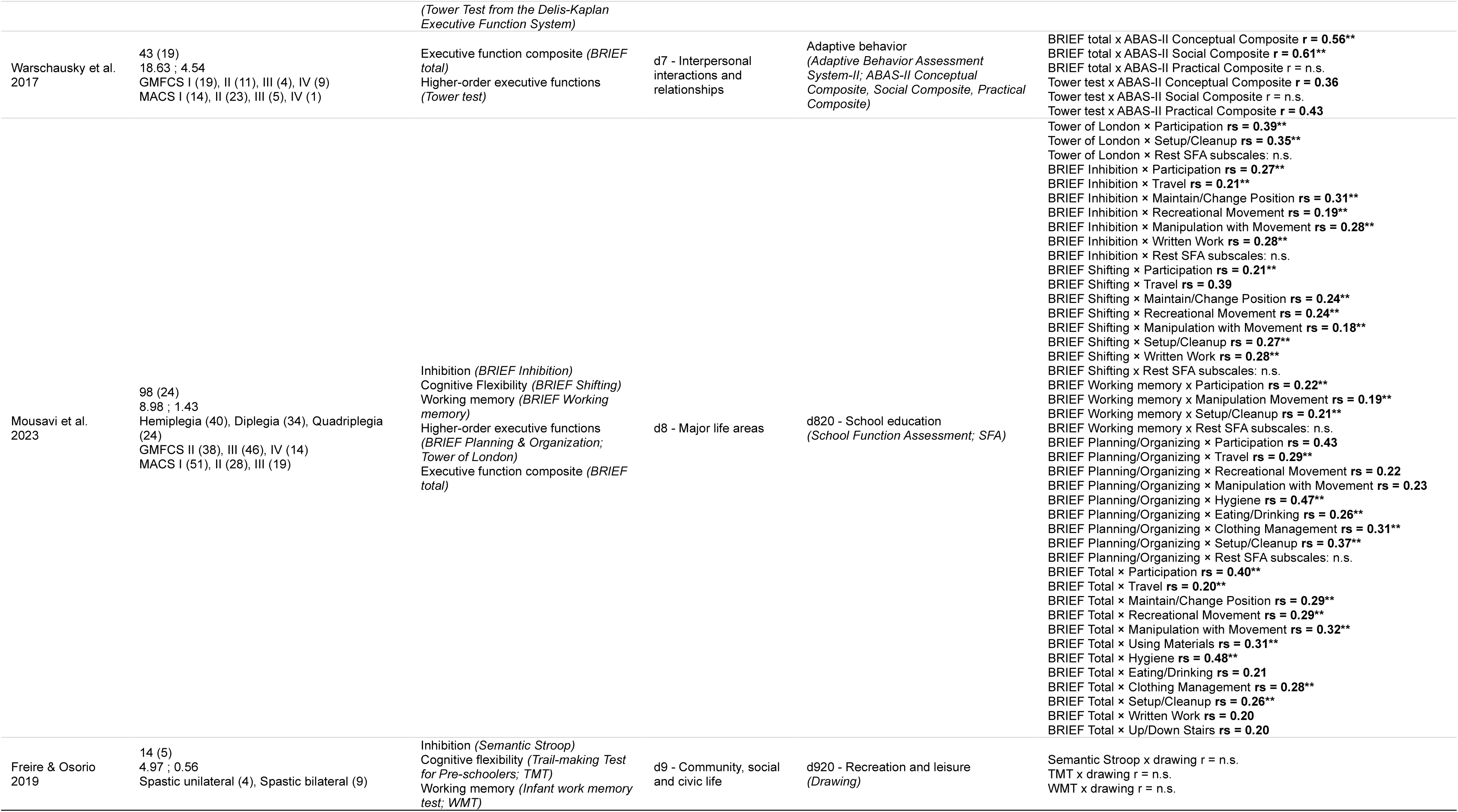
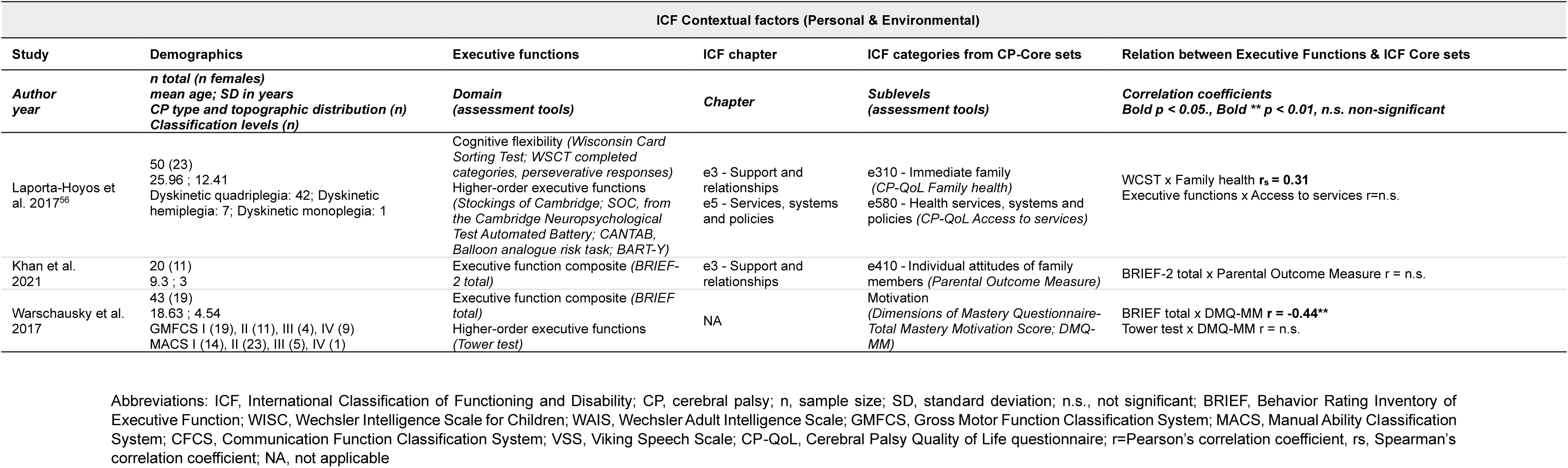
Summary of included study characteristics only for individuals with cerebral palsy.

| ICF Body Functions and Structures |  |  |  |  |  |
| --- | --- | --- | --- | --- | --- |
| Study | Demographics | Executive functions | ICF chapter | ICF categories from CP-Core sets | Relation between Executive Functions & ICF Core sets |
| Author year | *n* total (*n* females)<br>mean age; *SD* in years<br>CP type and topographic distribution (*n*)<br>Classification levels (*n*) | Domain<br>(assessment tools) | Chapter | Sublevels<br>(assessment tools) | Correlation coefficients<br>*Bold* $p < 0.05$., *Bold \*\** $p < 0.01$, *n.s.* non-significant |
| Stadskleiv et al. 2017 | 57 (38)<br>≈ 9.69 ; 2.92<br>Spastic hemiplegia (32); Spastic diplegia (16);<br>Spastic quadriplegia (4); Dyskinetic (5)<br>GMFCS I (36), II-III (11), IV-V (10)<br>MACS I (30), II-III (20), IV-V (7)<br>CFCS I-II (51), III (6)<br>VSS I-II (49), III-IV (8) | Executive function composite<br>(*Backward Memory from Leiter-R*,<br>*Wisconsin Card Sorting Test*; *WSCT*) | b1 - Mental functions | b117- Intellectual functions<br>(*Verbal Comprehension + Perceptual Reasoning Index = Cognitive quotient*) | Executive function composite x Cognitive quotient **$r = 0.58^{**}$** |
| Whittingham et al. 2014 | 46 (21)<br>11.1 ; 2.4<br>Spastic unilateral (46)<br>GMFCS I (35), II (11)<br>MACS I (6), II (40) | Executive function composite<br>(*BRIEF total*) | b1 - Mental functions | b117 – Intellectual functions<br>b140 – Attention functions<br>b152 – Emotion functions (*Strengths and Difficulties Questionnaire*; *SDQ*) | BRIEF total x *SDQ* Emotion $r = n.s.$<br>BRIEF total x *SDQ* Conduct **$r = -0.43^{**}$**<br>BRIEF total x *SDQ* Hyperactivity **$r = -0.49^{**}$**<br>BRIEF total x *SDQ* Peer $r = n.s.$<br>BRIEF total x *SDQ* Prosocial **$r = 0.28^{**}$** |
| van Rooijen et al. 2015 | 56 (19)<br>6 ; 0.58<br>Spastic unilateral (14); Spastic bilateral (28);<br>Dyskinetic (5); Spastic + Dyskinetic (4);<br>Dyskinetic + Ataxic (1)<br>GMFCS I (10), II (17), III (6), IV (8), V (7)<br>Unavailable (8) | Working memory<br>(*WISC-III Backward digit span*) | b1 - Mental functions | b117 - Intellectual functions<br>(*Raven Colored Progressive Matrices*)<br>b167 - Mental functions of language<br>(*Student Monitoring System for Language Performance*) | Digit span backward x *Raven Colored Progressive Matrices* **$r = 0.61^{**}$**<br>Digit span backward x *Student Monitoring System Language Performance* **$r = 0.40^{**}$** |
| van Rooijen et al. 2016 | 56 (19)<br>6 ; 0.58<br>Spastic unilateral (14); Spastic bilateral (28);<br>Dyskinetic (5); Spastic + Dyskinetic (4);<br>Dyskinetic + Ataxic (1)<br>GMFCS I (10), II (17), III (6), IV (8), V (7)<br>Unavailable (8) | Working memory (*Backward digit recall of Automated Working Memory Assessment*; *AWMA*) | b1 - Mental functions | b117 - Intellectual functions<br>(*Raven Colored Progressive Matrices*)<br>b167 - Mental functions of language<br>(*Dutch Language Proficiency Test*) | Backward digit recall *AWMA* x *Raven Colored Progressive Matrices* **$r = 0.52^{**}$**<br>Backward digit recall *AWMA* x *Dutch Language Proficiency Test* **$r = 0.48^{**}$** |
| Peeters et al. 2009 | 52 (19)<br>84 months (6.1)<br>Spastic hemiplegia (8); Spastic diplegia (22);<br>Spastic quadriplegia (12); Spastic-ataxic (5);<br>Spastic-hypotonic (1); Spastic-dyskinetic (1);<br>Ataxic (2)<br>MACS average 2.52 (SD=.94)<br>GMFCS average 2.71 (SD=1.26) | Working memory (*Forced-choice recognition task*) | b1 - Mental functions<br>b2 - Sensory functions and pain<br>b3 - Voice and speech functions | b117 - Intellectual functions (*Raven Coloured Progressive Matrices*)<br>b167 - Mental functions of language (*First-phoneme recognition task*)<br>b156 - Perceptual functions / b230 - Hearing functions(*Auditory Discrimination Task of the standardized Dutch Language Proficiency Test*)<br>b320 - Articulation functions (*Word Articulation task of the SLI Screening test*)<br>b330 - Speaking (*Timed word pronunciation*) | Forced-choice recognition task x *Raven Coloured Progressive Matrices* **$r = 0.48^{**}$**<br>Forced-choice recognition task x *First-phoneme recognition task* **$r = 0.57^{**}$**<br>Forced-choice recognition task x *Auditory Discrimination Task* **$r = 0.63^{**}$**<br>Forced-choice recognition task x *Word Articulation task* **$r = 0.73^{**}$**<br>Forced-choice recognition task x *Timed word pronunciation* **$r = 0.63^{**}$** |
| Li et al.<br>2014 | 44 (17)<br>10.40 ; 1.45<br>Spastic hemiplegia (18), Spastic diplegia (13),<br>Spastic-ataxic (2), Spastic-dyskinetic (1),<br>Athetoid (4), Ataxic (3) | Inhibition (*Day-Night Task*)<br>Cognitive flexibility (*Plus-Minus Task / Jersild Task*)<br>Working memory (*Running Memory Paradigm / Morris & Jones Task*) | b1 - Mental functions | b126 - Temperament and personality functions (*Theory of mind: False belief tasks; Faux pas tasks*) | Inhibition x False belief **r = -0.58\*\***<br>Inhibition x Faux pas **r = -0.37**<br>Information updating x False belief **r = 0.73\*\***<br>Information updating x Faux pas **r = 0.64\*\***<br>Shifting x False belief r = n.s.<br>Shifting x Faux pas r = n.s. |
| Caillies et al.<br>2012 | 10 (4)<br>9.3<br>Spastic bilateral (5); Spastic unilateral (4);<br>Ataxia (1) | Inhibition (*Stroop, Knock-tap test*)<br>Working memory (*Letter-Number Sequencing*) | b1 - Mental functions | b125 - Temperament and personality functions (*Theory of mind, Speaker's belief*) | Stroop x Theory of Mind r = n.s.<br>Stroop x Speaker's belief r = n.s.<br>Knock-tap test x Theory of mind r = n.s.<br>Knock-tap test x Speaker's belief r = n.s.<br>Letter-number seq x Theory of Mind r = n.s.<br>Letter-number seq x Speaker's belief Other-directed irony **r = 0.69** |
| Belmonte-Darraz et al. 2021 | 36<br>10.14 ; 2<br>13.3% walk independently<br>20% walk with assistive devices<br>66.7% reduced mobility in wheelchairs | Inhibition (*Emotion regulation checklist; ERC*) | b1 - Mental functions | b152 - Emotional functions (*Emotion matching task, Emotion situation knowledge, Expressive emotion knowledge, Receptive emotion knowledge*) | ERC x Emotion matching task r = n.s.<br>ERC x Emotion situation knowledge r = n.s.<br>ERC x Expressive emotion knowledge r = n.s.<br>ERC x Receptive emotion knowledge r = n.s. |
| Goble et al.<br>2012 | 11 (5)<br>33.0 ; 10.3<br>Spastic unilateral (3); Spastic bilateral (8)<br>GMFCS I (5), II (3), III (2), IV (1)<br>MACS I (3), II (8) | Working memory (*Backward Corsi block-tapping test*) | b1 - Mental functions<br>b2 – Sensory functions and pain | b163 - Basic cognitive functions<br>b260 - Proprioceptive function (*Ipsilateral Remembered position matching test*) | Backward spatial span x Variable error improvement **r = -0.67** |
| Laporta-Hoyos et al. 2017<sup>56</sup> | 50 (23)<br>25.96 ; 12.41<br>Dyskinetic quadriplegia: 42; Dyskinetic hemiplegia: 7; Dyskinetic monoplegia: 1 | Cognitive flexibility (*Wisconsin Card Sorting Test; WSCT completed categories, perseverative responses*)<br>Higher-order executive functions (*Stockings of Cambridge; SOC, from the Cambridge Neuropsychological Test Automated Battery; CANTAB, Balloon analogue risk task; BART-Y*) | b1 - Mental functions | b152 - Emotional functions (*CP-QoL Feelings about functioning*) | WCST x Feelings about functioning **r<sub>s</sub> = 0.51\*\***<br>WCST x Feelings about functioning **r<sub>s</sub> = 0.30** |
| Li et al.<br>2025 | 38 (15)<br>12 ; 2.96<br>Spastic (38)<br>GMFCS I (4), II (14), III (4), IV (7), V (9)<br>MACS II (13), III (12), IV (5), V (1), Unknown (1)<br>CFCS I (6), II (8), III (8), IV (7), V (9)<br>VSS I (8), II (7), III (10), IV (12), Unknown (1) | Executive function composite (*BRIEF-2 total*) | b1 - Mental functions | b152 - Emotional functions (*Developmental Behaviour Checklist – Primary Carer; DBC-P*) | BRIEF-2 total x DBC-P **r = 0.67\*\*** |
| Forsman & Eliasson 2016 | 46 (19)<br>11.01 ; 2.89<br>Unilateral spastic<br>MACS I (17), II (14), III (5) | Executive function composite (*Five To Fifteen parental questionnaire; FtF*) | b1 - Mental functions | b152 - Emotional functions<br>b156 - Perceptual functions<br>b167 - Mental functions of language (*Five To Fifteen parental questionnaire; FtF*) | FtF x Emotional/behavioral **r = 0.42\*\***<br>FtF x Language **r = 0.80\*\***<br>FtF x Memory **r = 0.75\*\***<br>FtF x Perception **r = 0.81\*\*** |
| Cabezas & Carriedo 2019 | 16 (6)<br>13.2 ; 4.20<br>Spastic (16)<br>GMFCS II (4), III (4), IV (8) | Inhibition<br>(*Anti-saccade task, Day-and-night task*) | b1 - Mental functions | b156 - Perceptual functions<br>(*Temporal processing*) | Anti-saccade task x Processing speed $r = \text{n.s.}$<br>Day-and-night x Processing speed reproduction $r = \mathbf{0.55}$<br>Day-and-night x Processing speed generalization $r = \mathbf{-0.60^{**}}$ |
| Soriano et al 2021 | 27 (12)<br>11.45 ; 0.77<br>Spastic hemiplegia (13); Spastic diplegia (5);<br>Spastic quadriplegia (4); Spastic unknown (2);<br>Ataxic (2); Unknown (1) | Executive function composite (*BRIEF total*) | b1 - Mental functions<br>b3 – Voice and speech functions | b140 - Attention functions (*Leiter International Performance Scale - Revised; Leiter-R*)<br>b167 - Mental functions of language<br>(*Test of Auditory Comprehension of Language, Fourth Edition ; TACL-4*)<br>b320 - Articulation functions<br>(*Test of Children's Speech Plus ; TOCS+*) | BRIEF total x TACL-4 $r = \text{n.s.}$<br>BRIEF total x Leiter-R $r = \text{n.s.}$<br>BRIEF total x TOCS+ $\rho = \text{n.s.}$ |
| Stadskleiv et al. 2014 | 29 (15)<br>11.35 ; 2.81<br>GMFCS I (1), III (2), IV (9), V (17)<br>MACS I (1), II (1), III (2), IV (8), V (17)<br>CFCS II (13), III (8), IV (2)<br>VSS III (1), IV (28) | Higher-order executive functions (*Be a Communicator Construction, Be a Communicator Description without naming*) | b1 - Mental functions | b167 - Mental functions of language<br>(*Test for the Reception of Grammar, second edition; TROG-2*) | Impulsivity x Language metrics $r = \text{n.s.}$<br>Planning x Language comprehension $r = \mathbf{0.40}$<br>Planning x Expressive verbal abilities $r = \mathbf{0.60^{**}}$<br>Monitoring x Language metrics $r = \text{n.s.}$ |
| Larsen et al. 2022 | 19<br>Unilateral (19)<br>Arterial ischemic stroke (16); Periventricular venous infarction (3) | Inhibition (*BRIEF Inhibition*)<br>Cognitive Flexibility (*BRIEF Shifting*)<br>Working memory (*BRIEF Working memory*)<br>Higher-order executive functions (*BRIEF Planning & Organization*)<br>Executive function composite (*BRIEF total*) | s1 - Structures of the nervous system | s110 - Structure of brain<br>(*Frontal white matter microstructure; Tractography*) | BRIEF Inhibition x White matter fractional anisotropy $rs = \text{n.s.}$<br>BRIEF Inhibition x White matter mean diffusivity $rs = \text{n.s.}$<br>BRIEF Inhibition x White matter axial diffusivity $rs = \mathbf{0.58}$<br>BRIEF Inhibition x White matter radial diffusivity $rs = \text{n.s.}$<br>BRIEF Shifting x White matter metrics $rs = \text{n.s.}$<br>BRIEF Working memory x White matter fractional anisotropy $rs = \text{n.s.}$<br>BRIEF Working memory x White matter mean diffusivity $rs = \mathbf{0.51}$<br>BRIEF Working memory x White matter axial diffusivity $rs = \mathbf{0.50}$<br>BRIEF Working memory x White matter radial diffusivity $rs = \text{n.s.}$<br>BRIEF Planning/Organizing x White matter metrics $rs = \text{n.s.}$<br>BRIEF total x White matter fractional anisotropy $rs = \text{n.s.}$<br>BRIEF total x White matter mean diffusivity $rs = \text{n.s.}$<br>BRIEF total x White matter axial diffusivity $rs = \mathbf{0.54}$<br>BRIEF total x White matter radial diffusivity $rs = \text{n.s.}$ |
| Di Lieto et al. 2017 | 19 (10)<br>8.7 ; 2.3<br>Spastic diplegia (19)<br>GMFCS I (2), II (10), III (3), IV (4)<br>MACS I (6), II (11), III (2) | Inhibition (*NEPSY-II Inhibition-inhibition speed*)<br>Cognitive flexibility (*NEPSY-II Inhibition-switching speed, NEPSY-II Response set, Inhibition-switching accuracy*) | s1 - Structures of the nervous system | s110 - Structure of brain<br>(*Neuroanatomical correlates; MRI*) | Inhibition, Cognitive flexibility domains x Corpus callosum score $r = \mathbf{-0.55}$<br>Inhibition, Cognitive flexibility domains x Other brain metrics $r = \text{n.s.}$ |
| Laporta-Hoyos et al. 2017<sup>55</sup> | 33 (15)<br>24.42 ; 12.61<br>Dyskinetic quadriplegia (28), Dyskinetic hemiplegia (4), Dyskinetic monoplegia (1)<br>GMFCS I (12), II (6), III (3), IV (5), V (7)<br>MACS I (5), II (8), III (11), IV (2), V (7)<br>CFCS I (14), II (13), III (2), IV (4) | Cognitive flexibility (*Wisconsin card sorting test; WCST*) | s1 - Structures of the nervous system | s110 - Structure of brain<br>(*Anatomical regions; MRI*) | WCST x Body & splenium of the corpus callosum (including the tapetum), bilateral posterior corona radiata, posterior thalamic radiation, retrolenticular part of the internal capsule, cingulum (posterior division) $rs = \mathbf{0.46}$<br>WCST x Right lateral occipital cortex (superior division), superior parietal lobe, precuneus cortex $rs = \mathbf{0.47}$<br>WCST x Right superior parietal lobe $rs = \mathbf{0.44}$<br>WCST x Right cingulate gyrus (anterior division) $rs = \mathbf{0.53}$ |
| Laporta-Hoyos et al. 2018 | 39 (19)<br>Median: 21 ; Interquartile range 13<br>Dyskinetic quadriplegia (32), Dyskinetic hemiplegia (6), Dyskinetic monoplegia (1) | Inhibition (*Stop signal task; SST*)<br>Cognitive flexibility<br>(*Wisconsin card sorting test; WCST, Lexical verbal fluency test*) | s1 - Structures of the nervous system | s110 - Structure of brain<br>(*Anatomical regions; MRI*) | SST x Parietal lobe $r = \mathbf{0.24}$<br>SST x Medial dorsal thalamus $r = \mathbf{-0.29}$<br>SST x rest anatomical regions $r = \text{n.s.}$<br>WCST x Global score $r = \mathbf{0.24}$ |
|  | GMFCS I (15), II (7), III (3), IV (5), V (9)<br>MACS I (5), II (10), III (12), IV (3), V (9)<br>CFCS I (15), II (16), III (4), IV (4), V (0) | Higher-order executive functions<br>(*Stockings of Cambridge test; SOC*) |  |  | WSCT x Frontal lobe **r = 0.34\*\***<br>WSCT x Parietal lobe **r = 0.25**<br>WSCT x Temporal lobe **r = 0.25**<br>WSCT x Posterior thalamus **r = 0.39\***<br>WSCT x Posterior limb of internal capsule **r = 0.26**<br>WSCT x Caudate **r = 0.29**<br>WSCT x Corpus callosum **r = 0.39\*\***<br>WSCT x rest anatomical regions **r = n.s.**<br>Lexical verbal fluency test x rest anatomical regions **r = n.s.**<br>SOC x Global score **r = -0.32\*\***<br>SOC x Frontal lobe **r = -0.32\*\***<br>SOC x Parietal lobe **r = -0.24**<br>SOC x Temporal lobe **r = -0.34\*\***<br>SOC x Occipital lobe **r = -0.29**<br>SOC x Medial dorsal thalamus **r = -0.37\*\***<br>SOC x Posterior thalamus **r = -0.25**<br>SOC x Caudate **r = -0.32\***<br>SOC x Corpus callosum **r = -0.41\*\***<br>SOC x rest anatomical regions **r = n.s.** |
| Pagnozzi et al. 2016 | 107 (57)<br>11.23 ; 2.83<br>Unilateral spastic (107) | Executive function composite (*BRIEF total*) | s1 - Structures of the nervous system | s110 - Structure of brain<br>(*Grey and white matter lesion burden correlates*) | BRIEF total x Lesion burden correlates **r = n.s.** |
| Meghji et al. 2024 | 61 (34)<br>12.56 ; 3.46<br>Unilateral spastic (61) | Executive function composite (*BRIEF total*) | s1 - Structures of the nervous system | s110 - Structure of brain<br>(*Functional connectivity within networks*) | BRIEF total x Functional connectivity **r = n.s.** after correction for multiple testing |
| da Rocha Dourado et al. 2013 | 76 (38)<br>8.9 ; 3.56<br>Spastic | Working memory (*Backward digit span, Backward Corsi block-tapping test*)<br>Higher-order EF (*Rey/Osterrieth Complex Figure task*) | s3 - Structures involved in voice and speech | s320 - Structure of mouth<br>(*Dental cavities*) | Backward Digit Span Test x number of teeth with cavities **r = -0.42\*\***<br>Complex Figure Test x number of teeth with cavities **r = -0.37\*\*** |

| ICF Activity |  |  |  |  |  |
| --- | --- | --- | --- | --- | --- |
| Study | Demographics | Executive functions | ICF chapter | ICF categories from CP-Core sets | Relation between Executive Functions & ICF Core sets |
| Author year | *n total (n females)*<br>*mean age; SD in years*<br>*CP type and topographic distribution (n)*<br>*Classification levels (n)* | *Domain*<br>*(assessment tools)* | *Chapter* | *Sublevels*<br>*(assessment tools)* | *Correlation coefficients*<br>*Bold p < 0.05., Bold \*\* p < 0.01, n.s. non-significant* |
| Jenks et al. 2007 | 57 (20)<br>7<br>Unilateral (22), Bilateral (32), Ataxic (3) | Working memory<br>(*Backward digits*) | d1 - Learning and applying knowledge | d155- Acquiring skills<br>d175 – Solving problems<br>(*Early numeracy test; ENT Number concept, Counting; Addition and subtraction test*) | Backward digits x ENT Number concept **r = 0.50\*\***<br>Backward digits x ENT Counting **r = 0.63\*\***<br>Backward digits x Addition **r = 0.65\*\***<br>Backward digits x Subtraction **r = 0.54\*\*** |
| Critten et al. 2024 | 15 (4)<br>9.06 ; 1.64<br>Spastic quadriplegia (7), Spastic hemiplegia (3), Diplegia (1), Athetoid-like (2), Other/unspecified (2) | Working memory<br>(*WMTB-C, Backward digit test*) | d1 - Learning and applying knowledge | d166 - Reading<br>(*One-minute reading, Nonsense passage reading*) | Backward digits x One-minute reading **r = 0.63**<br>Backward digits x Nonsense passage reading **r = 0.64\*\*** |
| Jenks et al.<br>2012 | 57 (20)<br>8.92 ; 0.69<br>Unilateral (22), Bilateral (32), Ataxic (3) | Inhibition (*Inhibition adapted by Jenks et al., 2009a*)<br>Cognitive flexibility (*Shifting adapted by Jenks et al., 2009a*)<br>Working memory (*Backward digits*) | d1 - Learning and<br>applying knowledge | d166 - Reading (*Three-Minute Reading test*)<br>d175 - Solving problems (*Student Monitoring System for Math's Achievement*) | Backward digits x Match achievement **r = 0.61\*\***<br>Backward digits x Reading **r = 0.59\*\*** |
| Jenks et al.<br>2009 | 57 (20)<br>7<br>Unilateral (22), Bilateral (32), Ataxic (3) | Inhibition (*Inhibition adapted by Jenks et al., 2009a*)<br>Cognitive flexibility (*Shifting adapted by Jenks et al., 2009a*)<br>Working memory (*Backward digits*) | d1 - Learning and<br>applying knowledge | d175 - Solving problems<br>(*Simple addition, Simple subtraction, Complex addition*) | Inhibition x Rest math metrics **r = n.s.**<br>Shifting x Simple addition reaction time **r = 0.28**<br>Shifting x Simple subtraction reaction time **r = 0.32\*\***<br>Shifting x Complex addition reaction time **r = 0.41\*\***<br>Shifting x Complex addition accuracy **r = -0.28**<br>Shifting x Rest match metrics **r = n.s.**<br>Backward digits x Simple addition reaction time **r = -0.49\*\***<br>Backward digits x Simple addition accuracy **r = 0.52\*\***<br>Backward digits x Simple subtraction reaction time **r = -0.45\*\***<br>Backward digits x Simple subtraction accuracy **r = 0.58\*\***<br>Backward digits x Complex addition reaction time **r = -0.39\*\***<br>Backward digits x Complex addition accuracy **r = 0.66\*\*** |
| Li et al.<br>2022 | 18 (10)<br>9.9 ; 3.1 | Inhibition (*NEPSY-II Auditory attention, Inhibition - Naming, Inhibition-Inhibition speed*)<br>Cognitive flexibility (*NEPSY-II Inhibition-switching, Animal sorting, Design fluency, Response set*) | d1 - Learning and<br>applying knowledge | d175 - Solving problems (*Calculation, Math fluency, Applied problems*) | Animal sorting x Match metrics **r = n.s.**<br>Auditory attention x Math metrics **r = n.s.**<br>Response set x Calculation **r = 0.57**<br>Response set x Rest math metrics **r = n.s.**<br>Design fluency x Math metrics **r = n.s.**<br>Inhibition-Naming x Math metrics **r = n.s.**<br>Inhibition-Inhibition x Applied problems **r = 0.75\*\***<br>Inhibition-Inhibition x Rest math metrics **r = n.s.**<br>Inhibition-Switching x Math metrics **r = n.s.** |
| Forsman &<br>Eliasson 2016 | 46 (19)<br>11.01 ; 2.89<br>Unilateral spastic<br>MACS I (17), II (14), III (5) | Executive function composite<br>(*Five To Fifteen parental questionnaire; FtF*) | d1 - Learning and<br>applying knowledge<br>d4 - Mobility | d155 - Acquiring skills<br>d440-d445 - Fine hand use, hand/arm use<br>d450-d455 - Walking, moving around<br>(*Five To Fifteen parental questionnaire; FtF*) | FtF x Learning **r = 0.76\*\***<br>FtF x Gross motor skills **r = 0.43\*\***<br>FtF x Fine motor skills **r = 0.70\*\*** |
| van Rooijen et al.<br>2012 | 116 (40)<br>7.3 ; 0.4<br>Spastic unilateral (42); Spastic bilateral (56);<br>Dyskinetic (14); Ataxic (4)<br>GMFCS I (56), II (20), III (17), IV (9), V (14) | Working memory (*Backward digit span WISC-III*) | d1 - Learning and<br>applying knowledge<br>d4 - Mobility | d175 - Solving problems (*Student Monitoring System for Arithmetic Performance*)<br>d440-d445 - Fine hand use, hand/arm use (*ABILHAND-Kids*)<br>d450-d455 - Walking, moving around<br>(*Gross Motor Function Measure-66; GMFM-66*) | Digit span backward x Student Monitoring System Arithmetic Performance **r = 0.37\*\***<br>Digit span backward x GMFM-66 **r = 0.40\*\***<br>Digit span backward x ABILHAND-Kids **r = 0.41\*\*** |
| van Rooijen et al.<br>2015 | 56 (19)<br>6 ; 0.58<br>Spastic unilateral (14); Spastic bilateral (28);<br>Dyskinetic (5); Spastic + Dyskinetic (4);<br>Dyskinetic + Ataxic (1)<br>GMFCS I (10), II (17), III (6), IV (8), V (7),<br>Unavailable (8) | Working memory (*Automated Working Memory Assessment; AWMA, Backwards digit recall*) | d1 - Learning and<br>applying knowledge<br>d4 - Mobility | d175 - Solving problems (*Early Numeracy Test-Revised; ENT-R, Counting*)<br>d440 - Fine hand use<br>d445 - Hand and arm use (*Box and Blocks test*) | Backward digit recall AWMA x ENT-R 6 years **r = 0.63\*\***<br>Backward digit recall AWMA x ENT-R 7 years **r = 0.66\*\***<br>Backward digit recall AWMA x Counting 6 years **r = 0.55\*\***<br>Backward digit recall AWMA x Counting 7 years **r = 0.63\*\***<br>Backward digit recall AWMA x Box and Blocks 6 years **r = 0.35**<br>Backward digit recall AWMA x Box and Blocks 7 years **r = 0.40** |
| van Rooijen et al. 2016 | 56 (19)<br>6 ; 0.58<br>Spastic unilateral (14); Spastic bilateral (28);<br>Dyskinetic (5); Spastic + Dyskinetic (4);<br>Dyskinetic + Ataxic (1)<br>GMFCS I (10), II (17), III (6), IV (8), V (7)<br>Unavailable (8) | Working memory (*Automated Working Memory Assessment; AWMA, Backwards digit recall*) | d1 - Learning and applying knowledge<br>d4 - Mobility | d175 - Solving problems (*Counting*)<br>d440-d445 - Fine hand use, hand/arm use (*Box and Blocks test*) | Backward digit recall AWMA x Box and Blocks test **r = 0.35\*\***<br>Backward digit recall AWMA x Counting **r = 0.55\*\*** |
| Ballester-Plané et al. 2018 | 52 (24)<br>24.9 (13)<br>Dyskinetic unilateral (8), Dyskinetic bilateral (44)<br>GMFCS I (15), II (8), III (6), IV (11), V (12)<br>BFMF I (7), II (12), III (16), IV (12), V (5)<br>MACS I (5), II (10), III (17), IV (10), V (10)<br>CFCS I (17), II (23), III (6), IV (6) | Cognitive flexibility (*Wisconsin Card Sorting Test; WSCT*) | d3 - Communication | d310 – Communicating with -receiving – spoken messages (*Communication Function Classification System; CFCS*) | WSCT x CFCS rs = n.s. |
| Pirila et al. 2004 | 15 (6)<br>Age range: 5-12<br>Spastic diplegia (15)<br>GMFCS I-III (9) IV-V (6) | Inhibition (*NEPSY Auditory Attention*)<br>Cognitive flexibility (*NEPSY Response Set*)<br>Higher-order executive functions (*NEPSY Tower test, NEPSY*) | d4 - Mobility | d410 - Changing basic body positions<br>d415 - Maintaining a body position<br>d420 - Transferring oneself (*Autti-Ramo's 1996 Scale; ARS*) | Executive function metrics x ARS r = n.s. |
| Kalkantzi et al. 2025 | 46 (20)<br>11.8 ; 2.8<br>Unilateral spastic (46)<br>MACS I (25) II (15) III (6) | Inhibition (*BRIEF Inhibition*)<br>Cognitive Flexibility (*BRIEF Shifting*)<br>Working memory (*BRIEF Working memory*)<br>Higher-order executive functions (*BRIEF Planning & Organization*)<br>Executive function composite (*BRIEF total*) | d4 - Mobility | d440 - Fine hand use,<br>d445 - Hand and arm use (*Assisting Hand Assessment; AHA, Children's Hand-Use Experience Questionnaire; CHEQ*) | BRIEF Inhibition x AHA **rs = -0.35**<br>BRIEF Inhibition x CHEQ grasp efficiency **rs = -0.34**<br>BRIEF Inhibition x CHEQ time **rs = -0.44**<br>BRIEF Inhibition x CHEQ bother **rs = -0.48**<br>BRIEF Shifting x CHEQ grasp efficiency **rs = -0.43**<br>BRIEF Shifting x CHEQ time **rs = -0.47**<br>BRIEF Shifting x CHEQ bother **rs = -0.50**<br>BRIEF Working memory x CHEQ grasp efficiency rs = n.s.<br>BRIEF Working memory x CHEQ time **rs = -0.41**<br>BRIEF Working memory x CHEQ bother **rs = -0.39**<br>BRIEF Planning x CHEQ grasp efficiency rs = n.s.<br>BRIEF Planning x CHEQ time **rs = -0.31**<br>BRIEF Planning x CHEQ bother **rs = -0.35**<br>BRIEF total x CHEQ grasp efficiency **rs = -0.42.**<br>BRIEF total x CHEQ time **rs = -0.47**<br>BRIEF total x CHEQ bother **rs = -0.48** |
| Warschawsky et al. 2017 | 43 (19)<br>18.63 ; 4.54<br>GMFCS I (19), II (11), III (4), IV (9)<br>MACS I (14), II (23), III (5), IV (1) | Executive function composite (*BRIEF total*)<br>Higher-order executive functions (*Tower test*) | d4 - Mobility | d440-d445 - Fine hand use, hand/arm use (*Manual Ability Classification System; MACS*)<br>d450-d455 - Walking, moving around (*Gross Motor Function Classification System; GMFCS*) | BRIEF total x MACS r = n.s.<br>BRIEF total x GMFCS r = n.s.<br>Tower test x MACS **r = -0.37**<br>Tower test x GMFCS r = n.s. |
| Soriano et al 2021 | 27 (12)<br>11.45 ; 0.77<br>Spastic hemiplegia (13); Spastic diplegia (5);<br>Spastic quadriplegia (4); Spastic unknown (2);<br>Ataxic (2); Unknown (1) | Executive function composite (*BRIEF total*) | d4 - Mobility | d440-d445 - Fine hand use, hand/arm use (*Manual Ability Classification System; MACS*)<br>d450-d455 - Walking, moving around (*Gross Motor Function Classification System; GMFCS*) | BRIEF total x MACS p = n.s.<br>BRIEF total x GMFCS p = n.s. |

| ICF Participation |  |  |  |  |  |
| --- | --- | --- | --- | --- | --- |
| Study | Demographics | Executive functions | ICF chapter | ICF categories from CP-Core sets | Relation between Executive Functions & ICF Core sets |
| Author year | *n total (n females)*<br>*mean age; SD in years*<br>*CP type and topographic distribution (n)*<br>*Classification levels (n)* | *Domain*<br>*(assessment tools)* | *Chapter* | *Sublevels*<br>*(assessment tools)* | *Correlation coefficients*<br>*Bold p < 0.05., Bold \*\* p < 0.01, n.s. non-significant* |
| Laporta-Hoyos et al. 2017<sup>56</sup> | 50 (23)<br>25.96 ; 12.41<br>Dyskinetic quadriplegia: 42; Dyskinetic hemiplegia: 7; Dyskinetic monoplegia: 1 | Cognitive flexibility (*Wisconsin Card Sorting Test; WCST*)<br>Higher-order executive functions (*Stockings of Cambridge; SOC, from the Cambridge Neuropsychological Test Automated Battery; CANTAB, Balloon analogue risk task; BART-Y*) | d3 - Communication<br>d8 - Major life areas<br>d9 - Community, social, civil life | d310 - Communicating with -receiving – spoken messages (*CP-QoL Communication and Physical health*)<br>d820 - School education (*CP QoL School wellbeing*)<br>d910-d920 - Engagement in social, recreational, and community life (*CP-QoL General well-being and participation*) | WCST x Communication & physical health **r = 0.36**<br>BART-Y x Communication & physical health **r = 0.31**<br>WCST x General wellbeing **r<sub>s</sub> = 0.36**<br>WCST x General wellbeing **r<sub>s</sub> = 0.40\*\***<br>BART-Y x General wellbeing **r<sub>s</sub> = 0.31**<br>Executive functions x School wellbeing r=n.s. |
| Nordberg et al. 2015 | 15 (7)<br>11.0 ; 1.4<br>Spastic unilateral (8), Spastic bilateral (2), Dyskinetic (2), Ataxic (3) | Working memory (*Backward Corsi block-tapping test; CBT, Backward digit span*) | d3 - Communication | d330-d360 - Speaking, Conversation, Using communication devices and techniques (*Test for Reception of Grammar-2; TROG-2, Peabody Picture Vocabulary Test - IV; PPVT-IV Bus Story Test; BST, Narrative Assessment Profile; NAP*) | CBT backward x Language metrics r = n.s.<br>Digit Span backward x Language metrics r = n.s<br>CBT backward x Storytelling metrics r = n.s. |
| Akyurek et al. 2024 | 22<br>Non-specified<br>Hemiplegia (22) | Executive function composite (*Executive Function and Occupational Routines Scale; EFORTS*) | d7 - Interpersonal interactions<br>d8 - Major life areas<br>d9 - Community social and civic life | d720 - Complex interpersonal interactions<br>d750 - Informal social relationships<br>d820 - School education<br>d920 - Recreation and leisure (*Child Occupation Self-Assessment Scale; COSA*) | EFORTS Morning-evening routines x Communication skills **r = 0.62**<br>EFORTS Morning-evening routines x Executive functions **r = 0.69\*\***<br>EFORTS Morning-evening routines x Motor skills **r = 0.53**<br>EFORTS Morning-evening routines x School functions **r = 0.71\*\***<br>EFORTS Morning-evening routines x Self-regulation **r = 0.63**<br>EFORTS Morning-evening routines x Instrumental Activities of Daily Living r = n.s.<br>EFORTS Morning-evening routines x Activities of Daily Living r = n.s.<br>EFORTS Play routine x Communication skills r = n.s.<br>EFORTS Play routine x Executive functions **r = 0.61**<br>EFORTS Play routine x Motor skills r = n.s.<br>EFORTS Play routine x School functions **r = 0.49**<br>EFORTS Play routine x Self-regulation **r = 0.49**<br>EFORTS Play routine x Instrumental Activities of Daily Living r = n.s.<br>EFORTS Play routine x Activities of Daily Living r = n.s.<br>EFORTS Social routine x Communication skills **r = 0.49**<br>EFORTS Social routine x Executive functions **r = 0.64**<br>EFORTS Social routine x Motor skills r = n.s.<br>EFORTS Social routine x School functions **r = 0.58**<br>EFORTS Social routine x Self-regulation **r = 0.55**<br>EFORTS Social routine x Instrumental Activities of Daily Living r = n.s.<br>EFORTS Social routine x Activities of Daily Living r = n.s. |
| Forsman & Eliasson 2016 | 46 (19)<br>11.01 ; 2.89<br>Unilateral spastic<br>MACS I (17), II (14), III (5) | Executive function composite (*Five To Fifteen parental questionnaire; FtF*) | d7 - Interpersonal interactions and relationships | d720 - Complex interpersonal interactions<br>d750 - Informal social relationships (*Five To Fifteen parental questionnaire; FtF*) | FtF x Social competency r = 0.64** |
| García-Galant et al 2024 | 60 (30)<br>10.15 ; 1.68<br>Spastic (54), Dyskinetic (5), Unknown (1)<br>Unilateral (36), Bilateral (24)<br>GMFCS I (34), II (18), III (6), IV (2)<br>MACS I (25), II (29), III (6)<br>BFMF I (32), II (20), III (7), IV (1)<br>CFCS I (36), II (19), III (3), IV (2) | <u>Core executive functions:</u><br>Inhibition (*NEPSY-II Inhibition index of Five Digit Test, Auditory Attention subtest*)<br>Cognitive flexibility (*NEPSY-II Response Set, Word Generation tasks*)<br>Working memory (*WISC-V Backward Digit Span, WNV Backward Spatial Span subtest*)<br><u>Higher-order executive functions</u> | d7 - Interpersonal interactions and relationships | d720 - Complex interpersonal interactions<br>d750 - Informal social relationships<br>d760 - Family relationships<br>d770 - Intimate relationships (*Affect Recognition, Theory of Mind NEPSY-II subtest*) | Core executive functions x Social cognition composite **rs = 0.47\*\***<br>Higher-order executive functions x Social cognition composite **rs = 0.37\*\***<br>Executive functions composite x Social Cognition composite **rs = 0.45\*\*** |
|  |  | (Tower Test from the Delis-Kaplan Executive Function System) |  |  |  |
| Warschawsky et al.<br>2017 | 43 (19)<br>18.63 ; 4.54<br>GMFCS I (19), II (11), III (4), IV (9)<br>MACS I (14), II (23), III (5), IV (1) | Executive function composite (BRIEF total)<br>Higher-order executive functions (Tower test) | d7 - Interpersonal interactions and relationships | Adaptive behavior (Adaptive Behavior Assessment System-II; ABAS-II Conceptual Composite, Social Composite, Practical Composite) | BRIEF total x ABAS-II Conceptual Composite **r = 0.56\*\***<br>BRIEF total x ABAS-II Social Composite **r = 0.61\*\***<br>BRIEF total x ABAS-II Practical Composite **r = n.s.**<br>Tower test x ABAS-II Conceptual Composite **r = 0.36**<br>Tower test x ABAS-II Social Composite **r = n.s.**<br>Tower test x ABAS-II Practical Composite **r = 0.43** |
| Mousavi et al.<br>2023 | 98 (24)<br>8.98 ; 1.43<br>Hemiplegia (40), Diplegia (34), Quadriplegia (24)<br>GMFCS II (38), III (46), IV (14)<br>MACS I (51), II (28), III (19) | Inhibition (BRIEF Inhibition)<br>Cognitive Flexibility (BRIEF Shifting)<br>Working memory (BRIEF Working memory)<br>Higher-order executive functions (BRIEF Planning & Organization; Tower of London)<br>Executive function composite (BRIEF total) | d8 - Major life areas | d820 - School education (School Function Assessment; SFA) | Tower of London x Participation **rs = 0.39\*\***<br>Tower of London x Setup/Cleanup **rs = 0.35\*\***<br>Tower of London x Rest SFA subscales: n.s.<br>BRIEF Inhibition x Participation **rs = 0.27\*\***<br>BRIEF Inhibition x Travel **rs = 0.21\*\***<br>BRIEF Inhibition x Maintain/Change Position **rs = 0.31\*\***<br>BRIEF Inhibition x Recreational Movement **rs = 0.19\*\***<br>BRIEF Inhibition x Manipulation with Movement **rs = 0.28\*\***<br>BRIEF Inhibition x Written Work **rs = 0.28\*\***<br>BRIEF Inhibition x Rest SFA subscales: n.s.<br>BRIEF Shifting x Participation **rs = 0.21\*\***<br>BRIEF Shifting x Travel **rs = 0.39**<br>BRIEF Shifting x Maintain/Change Position **rs = 0.24\*\***<br>BRIEF Shifting x Recreational Movement **rs = 0.24\*\***<br>BRIEF Shifting x Manipulation with Movement **rs = 0.18\*\***<br>BRIEF Shifting x Setup/Cleanup **rs = 0.27\*\***<br>BRIEF Shifting x Written Work **rs = 0.28\*\***<br>BRIEF Shifting x Rest SFA subscales: n.s.<br>BRIEF Working memory x Participation **rs = 0.22\*\***<br>BRIEF Working memory x Manipulation Movement **rs = 0.19\*\***<br>BRIEF Working memory x Setup/Cleanup **rs = 0.21\*\***<br>BRIEF Working memory x Rest SFA subscales: n.s.<br>BRIEF Planning/Organizing x Participation **rs = 0.43**<br>BRIEF Planning/Organizing x Travel **rs = 0.29\*\***<br>BRIEF Planning/Organizing x Recreational Movement **rs = 0.22**<br>BRIEF Planning/Organizing x Manipulation with Movement **rs = 0.23**<br>BRIEF Planning/Organizing x Hygiene **rs = 0.47\*\***<br>BRIEF Planning/Organizing x Eating/Drinking **rs = 0.26\*\***<br>BRIEF Planning/Organizing x Clothing Management **rs = 0.31\*\***<br>BRIEF Planning/Organizing x Setup/Cleanup **rs = 0.37\*\***<br>BRIEF Planning/Organizing x Rest SFA subscales: n.s.<br>BRIEF Total x Participation **rs = 0.40\*\***<br>BRIEF Total x Travel **rs = 0.20\*\***<br>BRIEF Total x Maintain/Change Position **rs = 0.29\*\***<br>BRIEF Total x Recreational Movement **rs = 0.29\*\***<br>BRIEF Total x Manipulation with Movement **rs = 0.32\*\***<br>BRIEF Total x Using Materials **rs = 0.31\*\***<br>BRIEF Total x Hygiene **rs = 0.48\*\***<br>BRIEF Total x Eating/Drinking **rs = 0.21**<br>BRIEF Total x Clothing Management **rs = 0.28\*\***<br>BRIEF Total x Setup/Cleanup **rs = 0.26\*\***<br>BRIEF Total x Written Work **rs = 0.20**<br>BRIEF Total x Up/Down Stairs **rs = 0.20** |
| Freire & Osorio<br>2019 | 14 (5)<br>4.97 ; 0.56<br>Spastic unilateral (4), Spastic bilateral (9) | Inhibition (Semantic Stroop)<br>Cognitive flexibility (Trail-making Test for Pre-schoolers; TMT)<br>Working memory (Infant work memory test; WMT) | d9 - Community, social and civic life | d920 - Recreation and leisure (Drawing) | Semantic Stroop x drawing **r = n.s.**<br>TMT x drawing **r = n.s.**<br>WMT x drawing **r = n.s.** |

| ICF Contextual factors (Personal & Environmental) |  |  |  |  |  |
| --- | --- | --- | --- | --- | --- |
| Study | Demographics | Executive functions | ICF chapter | ICF categories from CP-Core sets | Relation between Executive Functions & ICF Core sets |
| Author<br>year | *n* total (*n* females)<br>mean age; SD in years<br>CP type and topographic distribution (*n*)<br>Classification levels (*n*) | Domain<br>(assessment tools) | Chapter | Sublevels<br>(assessment tools) | Correlation coefficients<br>**Bold $p < 0.05$, Bold \*\* $p < 0.01$, n.s. non-significant** |
| Laporta-Hoyos et al. 2017<sup>56</sup> | 50 (23)<br>25.96 ; 12.41<br>Dyskinetic quadriplegia: 42; Dyskinetic hemiplegia: 7; Dyskinetic monoplegia: 1 | Cognitive flexibility (*Wisconsin Card Sorting Test; WSCT completed categories, perseverative responses*)<br>Higher-order executive functions (*Stockings of Cambridge; SOC, from the Cambridge Neuropsychological Test Automated Battery; CANTAB, Balloon analogue risk task; BART-Y*) | e3 - Support and relationships<br>e5 - Services, systems and policies | e310 - Immediate family (*CP-QoL Family health*)<br>e580 - Health services, systems and policies (*CP-QoL Access to services*) | WCST x Family health $r_s = \mathbf{0.31}$<br>Executive functions x Access to services $r = \text{n.s.}$ |
| Khan et al. 2021 | 20 (11)<br>9.3 ; 3 | Executive function composite (*BRIEF-2 total*) | e3 - Support and relationships | e410 - Individual attitudes of family members (*Parental Outcome Measure*) | BRIEF-2 total x Parental Outcome Measure $r = \text{n.s.}$ |
| Warschausky et al. 2017 | 43 (19)<br>18.63 ; 4.54<br>GMFCS I (19), II (11), III (4), IV (9)<br>MACS I (14), II (23), III (5), IV (1) | Executive function composite (*BRIEF total*)<br>Higher-order executive functions (*Tower test*) | NA | Motivation (*Dimensions of Mastery Questionnaire-Total Mastery Motivation Score; DMQ-MM*) | BRIEF total x DMQ-MM $r = \mathbf{-0.44^{**}}$<br>Tower test x DMQ-MM $r = \text{n.s.}$ |

Abbreviations: ICF, International Classification of Functioning and Disability; CP, cerebral palsy; n, sample size; SD, standard deviation; n.s., not significant; BRIEF, Behavior Rating Inventory of Executive Function; WISC, Wechsler Intelligence Scale for Children; WAIS, Wechsler Adult Intelligence Scale; GMFCS, Gross Motor Function Classification System; MACS, Manual Ability Classification System; CFCS, Communication Function Classification System; VSS, Viking Speech Scale; CP-QoL, Cerebral Palsy Quality of Life questionnaire; r=Pearson's correlation coefficient, $r_s$ , Spearman's correlation coefficient; NA, not applicable

### Quality assessment

Of the studies included, seven (18%) showed low, 20 (53%) moderate and 11 (29%) high RoB. Items within each QUIPS domain were not weighted; therefore, the RoB for each domain was determined based on the quantitative distribution of items addressed within that domain. The domains that demonstrated higher RoB were *Study participation* and *Study confounding*, as indicated in **Figure 3**.

**Figure 3.**
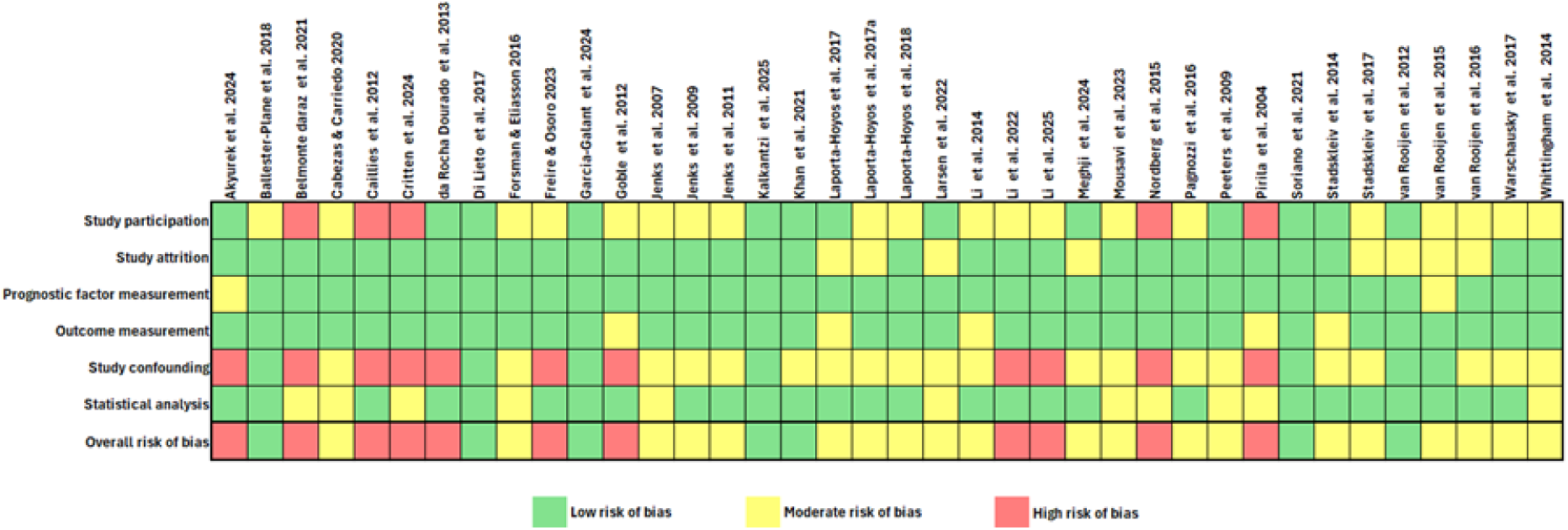
Study quality assessment using the Quality in Prognosis Studies (QUIPS) tool. Color coded: green represents low risk of bias, yellow represents moderate risk of bias, red represents high risk of bias.

Specifically, for the *Study participation* domain, 13 (34%) studies showed low, 20 (53%) moderate and 5 (13%) high RoB. This domain was affected mainly due to insufficient description of the sampling frame, period of recruitment and small sample. However, most studies reported the setting of recruitment, the in/exclusion criteria and provided a table with the baseline sample characteristics. Regarding *Study confounding*, seven (18%) of the included studies showed low, 20 (53%) moderate and 11 (29%) high RoB. Most of the studies did not identify or accounted for confounders either in their design or analyses, increasing the risk of various factors to mediate the relation between prognostic and outcome measure. The domains *Study attrition*, *Prognostic factor measurement* and *Outcome measurement* showed the lowest RoB. For *Study attrition*, 30 (79%) studies showed low and the remaining 8 (21%) moderate RoB. Attrition was mainly attributed to incomplete data due to comorbidities, indicating that the level of severity played a role in the completion of the included tests. Moderate RoB in the *Prognostic factor* and *Outcome measurement* was mainly attributed to absence of standardized/validated tests for the target population or test responses mediated by the examiner/parent when participant was unable to respond. It is worthwhile mentioning that in seven studies assessed as having an overall moderate RoB, the domains *Study confounding* and *Statistical analysis and reporting* were also judged to be at moderate risk. Although no domain was rated as high RoB in these studies, these moderate concerns in key methodological areas suggest the presence of potential bias and warrant cautious interpretation of the findings.

### Mean effect size

To quantify the magnitude of the relationship between EF and ICF domains, pooled ES from each study were calculated in three-level meta-analytic models. **Table 2** provides an overview of the ES characteristics included in each model.

**Table 2.**
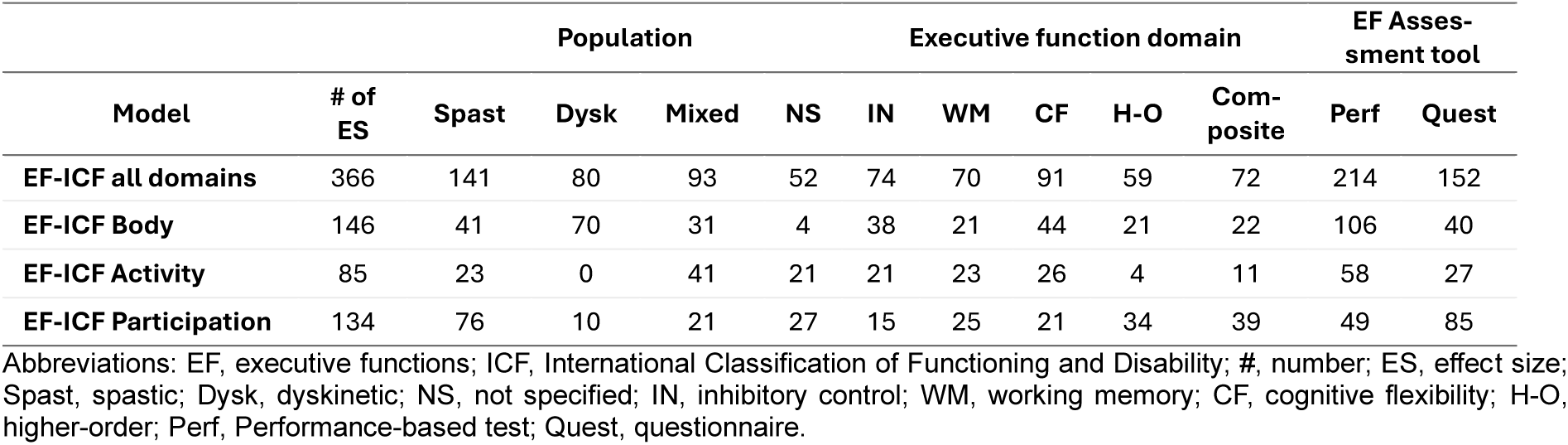
Frequency of effect sizes per model.

**Table 3** summarizes the three-level meta-analytic models across all domains. Overall, medium positive ES were found across all four unconditional models, indicating that better EF are related to better performance across ICF domains in people with CP. Specifically, a medium to large ES (r=0.26, p<0.001) was found between EF and ICF domains of Body Functions and Structures, Activity and Participation together. Looking at each ICF domain separately, a small to medium ES (r=0.21, p<0.01) was identified between EF and Body Functions and Structures and a medium to large ES between EF and Activity (r=0.38, p<0.001) and between EF and ICF Participation (r=0.26, p<0.01).

**Table 3.** Correlations between executive functions and ICF domains.

| Model | # of studies | # of ES | N | r | 95% CI | Q (df) | v level 1 (MSV) | v level 2 ($\sigma^2$) | v level 3 ($\sigma^3$) | I<sup>2</sup> |
| --- | --- | --- | --- | --- | --- | --- | --- | --- | --- | --- |
| EF-ICF all domains | 35 | 366 | 17.206 | 0.26*** | [0.15, 0.37] | 2134.27*** | 0.027 | 0.022 | 0.111 | 85.5% |
| EF-ICF Body | 20 | 146 | 4.817 | 0.21* | [0.00, 0.31] | 730.94*** | 0.033 | 0.023 | 0.178 | 85.7% |
| EF-ICF Activity | 11 | 85 | 3.537 | 0.38*** | [0.26, 0.49] | 191.57*** | 0.023 | 0.019 | 0.037 | 68.2% |
| EF-ICF Participation | 9 | 134 | 8.852 | 0.26** | [0.07, 0.43] | 709.32*** | 0.010 | 0.027 | 0.075 | 86.5% |

Abbreviations: EF, executive functions; ICF, International Classification of Functioning and Disability; #, number; ES, effect size; N, sample size included in the total number of effect sizes; r, correlation coefficient; CI, confidence interval; Q df, Q test for heterogeneity degrees of freedom; MSV, median sample variance; v, variance. Significance: \*\*\* $p<0.001$ , \*\* $p<0.01$ , \* $p<0.05$ .

### Publication bias

Results of the Egger’s test (**Table 4**) suggested evidence for publication bias in three out of the four models, specifically in the models EF-ICF all domains (p<0.001), EF-ICF Body Functions and Structures (p<0.001) and EF-ICF Participation (p<0.001).

**Table 4.** Results of the Egger’s test.

|  | Intercept | 95% CI | t | p-value |
| --- | --- | --- | --- | --- |
| EF-ICF all domains | -0.19 | [-0.30, -0.08] | 4.99 | <0.001*** |
| EF-ICF Body | 0.65 | [0.40, 0.90] | -4.27 | <0.001*** |
| EF-ICF Activity | 0.44 | [0.26, 0.63] | -0.79 | 0.427 |
| EF-ICF Participation | -0.56 | [-0.69, -0.44] | 8.04 | <0.001*** |

Abbreviations: EF, executive functions; ICF, International Classification of Functioning and Disability; CI, confidence interval. Significance: \*\*\* $p<0.001$ , \*\* $p<0.01$ , \* $p<0.05$

Funnel plots of these three models (**Figure S1**) also indicated asymmetries, probably due to small-study effects. The adjusted correlations of each model using the trim-and-fill method were as follows: EF-ICF all domains r=0.12, p<0.001, with the addition of 61 studies for symmetry restoration; EF-ICF Body Functions and Structures r=0.18, p<0.001, with the addition of 26 studies; EF-ICF Participation r=0.10, p<0.01, with the addition of 28 studies. Although coefficients relative to the originals were shrunk, they still remained robust, supporting the hypothesis that EF are related to ICF domains in people with CP.

### Sensitivity analyses

Sensitivity analyses excluding studies rated as high RoB yielded results comparable to the primary analyses (**Table 5**). Across all ICF domains, the pooled EF-ICF association remained statistically significant (r=0.26, p<0.001), with substantial heterogeneity (I²=88.1%). Significant associations were also observed for EF-ICF Activity (r=0.39, p<0.001, I²=75.4%). For EF-ICF Body Functions and Structures (r=0.22) and EF-ICF Participation (r=0.25), the pooled effect was similar in magnitude but did not reach statistical significance.

**Table 5.** Sensitivity analysis excluding high-risk-of-bias studies.

| Model | # of studies | # of ES | N | r | 95% CI | Q (df) | v level 1 (MSV) | v level 2 ($\sigma^2$) | v level 3 ($\sigma^2$) | $I^2$ |
| --- | --- | --- | --- | --- | --- | --- | --- | --- | --- | --- |
| EF-ICF all domains | 25 | 286 | 15754 | 0.26*** | [0.12, 0.40] | 1893.30*** | 0.023 | 0.023 | 0.112 | 88.1% |
| EF-ICF Body | 15 | 126 | 4352 | 0.22 | [-0.01, 0.45] | 664.69*** | 0.030 | 0.025 | 0.180 | 86.5% |
| EF-ICF Activity | 9 | 62 | 3228 | 0.39*** | [0.24, 0.55] | 174.42*** | 0.021 | 0.022 | 0.041 | 75.4% |
| EF-ICF Participation | 6 | 98 | 8174 | 0.25 | [-0.02, 0.54] | 510.71*** | 0.011 | 0.026 | 0.101 | 91.3% |

Abbreviations: EF, executive functions; ICF, International Classification of Functioning and Disability; #, number; ES, effect size; N, sample size included in the total number of effect sizes; r, correlation coefficient; CI, confidence interval; Q df, Q test for heterogeneity degrees of freedom; MSV, median sample variance; v, variance. Significance: \*\*\* $p < 0.001$ , \*\* $p < 0.01$ , \* $p < 0.05$ .

### Moderator analyses

Heterogeneity across the three-level meta-analytic models was statistically significant, as indicated by significant Q statistics in **Table 3**. The model examining the relationship between EF and ICF Activity showed moderate heterogeneity (I²=68.2%), whereas the models examining EF in relation to all ICF domains combined, Body Functions and Structures, and Participation demonstrated substantial heterogeneity (I²>75%).^74^ This indicates considerable variability in ES across studies. Consequently, moderator analyses were conducted to examine whether selected study characteristics (i.e., EF domain, CP subtype, type of EF assessment tool, mean age) accounted for variability in the relationship between EF and all ICF domains combined.

Results of the moderator analyses are presented in **Table 6**. EF domain did not significantly moderate the relationship between EF and all ICF domains combined (Wald test, p=0.492). Nevertheless, pooled ES estimates varied descriptively across EF domains. Estimates were largest for working memory (r=0.39, p<0.001), followed by cognitive flexibility (r=0.25, p<0.001), higher-order EF (r=0.24, p=0.036) and inhibitory control (r=0.21, p=0.001). Smaller, non-significant pooled ES were observed for EF composite measures (r=0.18). CP subtype significantly moderated the relationship between EF and all ICF domains (Wald test, p=0.023). Studies including mixed CP samples yielded a large and statistically significant pooled ES (r=0.47, p<0.001), indicating that the strength of the EF-ICF relationship differed according to CP subtype, particularly in mixed samples. In contrast, pooled estimates for spastic (r=0.14) and dyskinetic (r=−0.16) CP subtypes were smaller and not statistically significant. The type of EF assessment tool was also not a significant moderator of the relationship between EF and the combined ICF domains (Wald test, p=0.189). Descriptively, studies using performance-based EF measures showed a moderate and statistically significant pooled ES (r=0.33, p<0.001), whereas studies using questionnaire-based measures showed a smaller, non-significant pooled ES (r=0.13). Finally, participants’ mean age significantly moderated the relationship between EF and ICF domains (z=−0.03, p=0.013), such that studies with older samples demonstrated smaller pooled ES.

**Table 6.** Results of moderation analysis.

| Categorical moderators |  |  |  |  |  |  |
| --- | --- | --- | --- | --- | --- | --- |
| Moderator<br>(*p*-value of omnibus<br>Wald's) | Levels | *z* (SE) | *r* [95% CI] | *t* | *df* | *p*-value |
| **Executive function domain**<br>(0.492) | Inhibitory control | 0.21 (0.07) | 0.21 [0.07, 0.33] | 2.84 | 23.8 | **0.001\*\*** |
|  | Working memory | 0.39 (0.08) | 0.37 [0.23, 0.50] | 5.12 | 24.5 | **<0.001\*\*\*** |
|  | Cognitive flexibility | 0.25 (0.06) | 0.24 [0.13, 0.35] | 3.77 | 24.5 | **<0.001\*\*\*** |
|  | Higher-order EF | 0.24 (0.11) | 0.24 [0.02, 0.43] | 2.25 | 20 | **0.036\*** |
|  | EF composite | 0.18 (0.09) | 0.18 [0.00, 0.34] | 2.05 | 17.9 | 0.055 |
| **Cerebral palsy subtype**<br>(0.023) | Spastic | 0.14 (0.13) | 0.14 [-0.11, 0.38] | 1.06 | 10.19 | 0.313 |
|  | Dyskinetic | -0.16 (0.16) | -0.16 [-0.44, 0.15] | -0.98 | 1.99 | 0.431 |
|  | Mixed | 0.47 (0.04) | 0.44 [0.37, 0.50] | 10.6 | 14.79 | **<0.001\*\*\*** |
| **Assessment tool**<br>(0.189) | Performance-based | 0.33 (0.06) | 0.32 [0.21, 0.42] | 5.52 | 30.4 | **<0.001\*\*\*** |
|  | Questionnaire | 0.13 (0.09) | 0.13 [-0.05, 0.30] | 1.34 | 12.9 | 0.202 |
| Continuous moderators |  |  |  |  |  |  |
| Moderator |  | *z* (SE) | / | *t* | *df* | *p*-value |
| **Age** | Average mean age | -0.03 (0.01) | / | -3.49 | 5.80 | **0.013\*** |

Abbreviations: EF, executive functions; SR, standard error; ES, effect size; *df*, degrees of freedom. Significance: \*\*\* $p<0.001$ , \*\* $p<0.01$ , \* $p<0.05$ .

## DISCUSSION

This is, to the authors’ knowledge, the first systematic review and meta-analysis to examine the relationship between EF and all domains of functioning within the ICF framework in individuals with CP. ICF components were classified using the CP ICF Core Sets to ensure relevance for the target population. A narrative synthesis was conducted to provide a comprehensive overview of the existing literature on the association between EF and ICF domains, while meta-analyses were used to quantify the strength of these associations and to identify influential factors.

### Descriptive characteristics

Across the 38 included studies, participant characteristics were highly heterogeneous, in terms of age, motor disorder and topographic subtype, as well as functioning. Twenty-eight studies focused on children with mean age between 7-12 years, a period that represents a critical developmental window during which EF undergo substantial refinement.^75^ This age range also coincides with formal schooling, when increasing academic and behavioral demands place greater reliance on EF, making them particularly important for learning, classroom functioning, and educational support needs.^75^ However, studies including adults with CP were underrepresented, in line with findings of previous reviews.^21,76^ This highlights the emphasis on childhood in research on childhood-onset disabilities, despite the fact that these children transition into adulthood.^77,78^ Regarding CP subtype, eight studies (21%) did not specify either the type of motor disorder and 12 (31.5%) the topographic distribution, making classification challenging. Additionally, absence of this information hinders obtaining knowledge with a higher prognostic value. About 39% of the included studies examined mixed CP motor types, which increases generalization but reduces specificity to distinct clinical presentations, as impairments can differ substantially between subtypes, such as spastic and dyskinetic CP. About 26% of the participants with reported data were classified as GMFCS IV and V, reflecting inclusion of cohorts with severe motor function. MACS was skewed toward milder levels, with levels I-III adding up to about 88%, consistent with over-representation of unilateral spastic CP.^79^ Lastly, despite the prevalence of ADHD and ASD in CP^80^ and their well-established association with EF impairments,^81^ only a limited number of studies reported their co-occurrence. The absence of such reporting or its incorporation into statistical analysis can lead to confounding, as it becomes unclear whether observed EF difficulties are core features of CP and/or the neurodevelopmental condition.

Delving into the assessment of EF, the lack of established and standardized protocols for assessing cognitive function in individuals with CP^82^ was evident in the variability of conceptual frameworks used to operationalize EF across studies. EF constructs were assessed inconsistently, with no systematic pattern according to CP subtype or outcome measure. Working memory was the most frequently assessed construct (18 studies), followed by inhibitory control (15 studies) and cognitive flexibility (15 studies). The particular emphasis on working memory reflects the extensive evidence on the importance of this construct in academic performance^75,83^ and the subsequent focus on school-aged children. Assessment tools were likewise heterogeneous, with many instruments originally developed for typically developing populations and assuming intact motor, speech, and visual-perceptual function. In the context of CP, these commonly co-occurring impairments can affect test outcomes, particularly in individuals with more severe disabilities, as highlighted by Fluss and Lidzba.^84^ This challenge was further reflected in study attrition, as inability to complete EF tasks was frequently marked as missing data in the QUIPS checklist. Therefore, appropriate test accommodations and alternative ways to assess cognition ^85,86^ are essential.

### Descriptive relations between EF and ICF domains

A gradient was observed in the distribution of studies across ICF domains, with the largest number focusing on Body Functions and Structures, followed by progressively fewer studies addressing Activity, Participation, and Contextual factors.

Regarding the ICF Body Functions and Structures domain, most studies targeted the relation between EF and the ICF Mental functions chapter (b1), particularly intellectual, language and emotional function. Results suggested that better EF were related with stronger mental functions across either CP subtype. This focus is expected since EF constructs are themselves embedded within this ICF chapter. Consequently, much of the existing literature aims to disentangle the complex interrelationships among cognitive domains in order to delineate more accurate cognitive profiles.^84,87^ Another well-studied ICF chapter in relation to EF was Structure of the nervous system (s1), reflecting the effort to identify neuroanatomical correlates of EF impairments to support earlier and more accurate prognosis. Across studies, brain lesion characteristics showed small to moderate associations mainly with working memory and cognitive flexibility, whereas relationships with EF composite scores were weaker and less consistent. This might be explained by the fact that EF rely on a widespread neural network, whereby individual EF components are supported by partially distinct neural substrates.^88^ Therefore, EF composite scores may be less sensitive to specific structure-function relationships than distinct EF components.

At the level of Activity, most studies focused on the ICF chapters Learning and applying knowledge (d1) and Mobility (d4). Within the d1 chapter, studies targeted school-aged children with CP and explored mathematics and reading in relation to working memory mainly, given its key role in learning processes.^89,90^ Correlations were primarily moderate, indicating that better working memory and EF in general are associated with stronger academic outcomes. Mobility was mainly represented by the GMFCS, the MACS and upper limb performance-based tests. Particularly metrics of the upper limb showed small to moderate relations with all EF constructs in children with spastic CP and good functional level. The EF associations with upper limb function may suggest that fine motor skills require increased cognitive engagement, relying on more advanced cognitive processes.^14,91^ At the same time, alternative explanations should be considered. First, the tasks used to assess fine motor skills may inherently recruit additional cognitive domains which might be impaired. Second, poor upper limb function may limit the ability to adequately perform tasks used to assess EF, particularly those requiring manual responses. In this case, lower EF scores may partially reflect task performance constraints rather than true deficits in EF. Finally, it is possible that these associations arise from the shared neural substrates, whereby the underlying lesion simultaneously affects both EF and motor function Therefore, the relationship between EF and upper limb function is likely multifactorial, reflecting both the cognitive demands inherent to fine motor function^92^ and the potential impact of motor limitations on test performance. Absence of EF-mobility associations in more severe CP subtypes (e.g., dyskinetic) in the present study likely reflects the lack of assessment tools that can reliably capture cognitive and upper limb function in people with severe disability.^93^

ICF Participation was mainly expressed through the chapter Interpersonal interactions and relationships (d7), followed by Major life areas (d8) and Community, social and civic life (d9), suggesting an interest in understanding how EF contribute to independence and engagement in social life.^87^ As expected, most components were assessed through questionnaires. Although studies targeting participation were sparse, the number of reported correlations was relatively high, probably due to the many items included in questionnaires. Here again, small to moderate correlations were shown between EF and the corresponding ICF chapters in spastic or mixed CP populations.

Only three studies examined ICF Contextual Factors in relation to EF, consistent with underrepresentation of this domain as found by Carracedo-Martín et al.^21^ The few correlations were small, indicating a positive significant relation between EF and context in adults with CP. The limited representation of contextual factors in the present review likely reflects broader trends within this research area. In CP research, priorities have traditionally centered on neurological impairment and its subsequent motor and rest functional outcomes. This clinical emphasis may contribute to the relative neglect of contextual factors, which are often viewed as secondary influences. Moreover, EF research in CP has predominantly focused on body- and activity-level, with contextual influences being less frequently conceptualized as important determinants, despite their central role within the ICF framework. In contrast, research in typically developing children consistently identifies contextual factors, including socioeconomic status, parental education, emotional support, and classroom management, as important EF moderators.^83,94,95^ Although recent CP research has begun to show a promising shift toward greater consideration of contextual factors,^96,97^ this shift is yet to be fully reflected in empirical studies examining EF.

### Quantitative relations between EF and ICF domains

Unconditional three-level meta-analytic models revealed significant positive associations between EF and all ICF domains. The grand model associating EF with all ICF domains yielded a medium to large ES (r=0.26), suggesting that EF indeed contribute to multiple aspects of daily functioning in individuals with CP.

Across domains, ICF Activity demonstrated the strongest association with EF, characterized by the largest ES (r=0.38), the narrowest confidence interval, and the lowest heterogeneity. This finding aligns with previous work suggesting that EF are essential for task execution.^98,99^ In CP, where motor impairments increase the cognitive demands of everyday actions, successful activity execution may rely more on EF to compensate for motor inefficiencies.^100,101^ Alternatively, poorer EF outcomes in people with CP might reflect the unadjusted nature of EF assessment tasks for this population. Associations between EF and ICF Body Functions and Structures yielded a medium to large ES (r=0.21), likely reflecting the embeddedness of EF within broader mental functions while preserving their role as higher-order control processes, separate from other cognitive domains. Likewise, the association between EF and ICF Participation domain showed a medium to large ES (r=0.26), underscoring the broad relevance of EF beyond task execution alone.

A theory-driven reason that might explain the strength of these associations is the following. Execution of activities requires moment-to-moment regulation of behavior.^7^ EF are continuously essential for maintaining task goals, monitoring performance, correcting errors, and adjusting strategies in real time,^5,7^ thereby reflecting a strongest relationship with measures at the level of activity. In contrast, Body Functions and Structures represent underlying capacities and systems rather than ongoing goal-directed behavior. As such, they might not consistently demand moment-to-moment behavioral regulation in the same way that activity does.^102^ Likewise, participation outcomes often reflect long-term involvement or contextual opportunity. Here, EF drive self-regulation and adaptation while interacting with external factors.^103^ In addition, outcome measures within the ICF Activity domain were relatively homogeneous, which may have contributed to the stability of the associations. For instance, three studies by Jenks et al.^51–53^ and three by van Rooijen et al.^70–72^ used overlapping measures of reading and arithmetic performance, thereby reducing variability across academic outcomes. Similarly, upper limb activity was assessed using well-established and widely used instruments in CP (i.e., Box and Blocks Test, Assisting Hand Assessment, ABILHAND-Kids, Children’s Hand-Use Experience Questionnaire, MACS). The use of comparable, population-specific and standardized measures likely enhanced consistency across studies and strengthened the observed EF-ICF Activity associations. Nevertheless, although ICF Activity shows the strongest association due to its immediate reliance on EF, the significant medium to large ES across the other ICF domains collectively underscore the important role of EF in shaping functioning in CP.

ES further varied depending on analytical decisions. Sensitivity analyses, including adjustments for publication bias and high RoB studies, produced more conservative estimates in some domains. To account for publication bias, the trill-and-fill method restored the funnel plot asymmetry and readjusted the ES^104^ to r=0.12 for EF-ICF all domains, r=0.18 for EF-ICF Body Functions and Structures and r=0.10 for EF-ICF Participation. Given that heterogeneity and outcome diversity can further contribute to funnel plot asymmetry,^105^ the trim-and-fill-adjusted estimates shall be best interpreted as conservative lower-bound estimates rather than representations of the true associations. Despite this attenuation, the overall pattern of findings remained largely consistent, with ES continuing to indicate significant positive associations between EF and ICF domains, supporting the primary results. Further sensitivity analyses excluding high RoB studies yielded ES comparable to the original estimates across domains, with the exception of ICF Body Functions and Structures and Participation. Although these models retained a medium ES, they did not reach statistical significance. These results should also be interpreted cautiously, as the sensitivity analysis for ICF Participation especially was likely underpowered, including only six studies.^106^ Overall, the persistence of medium to medium-large ES after exclusion of high RoB studies indicates that the observed associations are not driven by methodological weaknesses and remain rather stable, underscoring the meaningful contribution of EF to functioning in CP.

### Moderator analyses

Substantial heterogeneity was observed at both between-study and within-study levels, justifying the use of three-level random-effects models. Therefore, moderator analyses of the grand model (i.e., ICF all domains) was used to refine the interpretation of the findings. As hypothesized, CP subtype significantly moderated the association between EF and all ICF domains, with mixed CP samples showing the strongest effects, while associations were weaker and non-significant in spastic and dyskinetic subtypes when examined separately. This finding might be explained by the fact that mixed samples may capture greater variability in motor and cognitive profiles, increasing the contribution of EF to overall functioning.^87,107^ In contrast, variability was considerably lower particularly in dyskinetic CP, whereby data was stemming -with some variation-from repeated samples. This further underlines the scarcity of studies in this population and emphasizes the absence of validated access systems and focus on severe cases.^84^ Age also emerged as a significant moderator, with increasing age associated with slightly weaker EF-ICF associations.^107,108^ One possible explanation is the protracted developmental trajectory of EF, which reach maturity slower, when activity and participation might already be stabilized, consequently producing weaker observed EF-ICF association with age. Additionally, the development of routines and systematic habits with age^108^ may reduce reliance on active behavioral regulation.

### Research and clinical implications

The structured mapping of the existing literature, together with the identification of patterns and inconsistencies, highlights several important avenues for future research. First, there is a clear need to develop and adopt standardized evaluation protocols for assessing EF in CP, which would enhance methodological consistency, improve comparability across studies, and strengthen the utility of research findings for clinical decision-making. Standardization of EF assessment lies both in the unified use of a framework as well as specific tools that assess each EF construct embedded in the framework. Second, alternative and reliable methods for assessing EF in individuals with severe disabilities are essential to ensure representation across the full spectrum of CP. In this regard, accessible methods, developed by Stadskleiv et al.^69,86^ and Bekteshi et al.,^85^ to assess people with severe motor and speech impairment are promising and warrant further validation in larger cohorts. Third, EF should be more systematically examined within ICF Contextual factors, which remain underrepresented despite their relevance to functioning. Furthermore, EF research should also be directed towards adults with CP and potentially explore how mature EF can assist in the adoption of routines. Finally, given the high RoB identified using the QUIPS and the multifactorial nature of EF emphasized by Best and Miller,^109^ future studies should more rigorously address potential confounding in appropriate study design, analysis and clear reporting.

These findings also have direct implications for clinical practice. The multiple associations of EF with multiple domains of functioning, emphasize that EF assessment should be incorporated into routine clinical practice for individuals with CP. The stronger and more consistent associations between EF and ICF Activity further support the clinical relevance of EF assessment. Here, activity outcomes may serve as a particularly sensitive domain for evaluating the effect of EF-focused interventions in CP. From a practical point, these results also support interventions targeting EF not only as a means of improving complex cognition, but also as a way to enhance functional outcomes that align with the ICF framework and the corresponding Core Sets. The moderate associations for Body Functions and Structures and Participation further support this view, suggesting that EF contribute to functioning through multiple pathways, including regulation of underlying systems and adaptation to social and environmental demands. Although our results do not establish causality, the observed associations support considering that EF-focused interventions may be more beneficial when embedded within meaningful daily activities, rather than trained in isolation. Finally, the moderating age effect can inform interventions in placing more emphasis on EF enhancement in younger children, while focusing on strategy optimization, consolidation of routines, and environmental supports when it comes to older people.

### Considerations

A few points warrant further consideration. First, mapping outcomes into ICF domains required theoretical judgment, as most studies did not explicitly frame their measures within the ICF framework. Although classification decisions were guided by established ICF definitions, some degree of subjectivity is probably present. Second, the uneven distribution of outcomes across ICF domains limits the precision of some domain-specific estimates. In the same vein, outcomes within ICF domains were heterogeneous, spanning from emotions to dental carries. This heterogeneity complicates the interpretation of pooled ES, as outcomes grouped under the same ICF domain reflect qualitatively different aspects of functioning. Nevertheless, running meta-analytic models on distinct ICF chapters was not possible, due to the reduced power resulting from the modest number of included studies. Similarly, multiple moderator analyses could also reduce the importance of some of the current single moderators due to insufficient power. The current meta-analysis provides a view of the global relations between EF and ICF domains, while moderator analyses aid the understanding of factors influencing this relation.

## CONCLUSION

Although typically viewed as a domain of cognitive function, EF have a meaningful association with all domains of daily functioning in people with CP. Findings from the 38 included studies, together with the pooled estimates from the present meta-analyses, indicate that EF are moderately associated with a range of constructs across ICF domains. These associations suggest that EF impairments may be reflected not only in cognitive function but also in aspects of functioning that extend beyond cognition. Conversely, better EF performance may be linked to more favorable functioning across other domains, highlighting the potential relevance of EF as a cross-cutting component of everyday functioning in CP and a meaningful intervention target. Marked heterogeneity in study characteristics supports the need for standardized EF assessment protocols in CP, including standard mapping of EF constructs and the use of common, well-validated assessment tools to enhance comparability across studies. At the same time, the current literature base remains skewed toward individuals with milder motor impairments, while adults are more rarely included. The development and further validation of accessible EF assessment methods for individuals with severe disabilities is therefore essential to ensure representation of the full CP spectrum and to avoid the systematic exclusion of those with the greatest support needs.

## ACKNOWLEDGMENTS

The authors declare no interests that might be perceived as conflict or bias. Alexandra Kalkantzi as first author was supported by funding from the European Commission, Horizon Europe Research and Innovation Action under Grant Agreement No 101057309, in the context of the AINCP project. Saranda Bekteshi as last author was supported by FWO junior postdoctoral fellowship, project nr 1264123N. The work for Giuseppina Sgandurra was also partially supported by the Italian Ministry of Health Grant RC2025 (and the 5 ×1000 voluntary contributions). The funders had no role in the design of the systematic review or in the drafting of the review protocol or manuscript. The authors acknowledge the use of ChatGPT (OpenAI, San Francisco, CA, USA) to assist with language editing (i.e., provision of synonyms, sentence structure improvement).

## DATA AVAILABILITY STATEMENT

The data that support the findings of this study are available from the corresponding author upon reasonable request.

