## Supplementary material for "Executive Functions and ICF Core Sets in Cerebral Palsy: A Systematic Review and Meta-Analysis"

**Table S1.** Search strategy, tailored to each database, and corresponding results

| Database | Search string; Search conducted in 14 July 2025 | Results |
|---|---|---|
| PubMed | (("cerebral palsy"[All Fields] OR ("perinatal"[All Fields] AND "stroke"[All Fields])) AND ("relat*"[All Fields] OR "associat*"[All Fields] OR "correlat*"[All Fields]) AND ("executive function"[All Fields] OR "executive control"[All Fields] OR "self-regulation"[All Fields] OR "cognition"[MeSH Terms] OR "cognition"[All Fields] OR ("executive"[All Fields] AND "control"[All Fields]) OR ("self-control"[MeSH Terms] OR "self-control"[All Fields] OR ("self"[All Fields] AND "regulation"[All Fields]) OR "self-regulation"[All Fields]) OR "working memory"[All Fields] OR "inhibition"[All Fields] OR ("inhibitory"[All Fields] AND "control"[All Fields]) OR "cognitive flexibility"[All Fields])) | 870 |
| EMBASE | ('cerebral palsy' OR 'cerebral palsy' OR ('perinatal' AND 'stroke')) AND ('relation*':ti,ab,kw OR 'associat*':ti,ab,kw OR 'correlat*':ti,ab,kw) AND ('executive function'/exp OR 'executive function' OR 'self-regulation' OR 'cognition' OR 'cognition' OR 'executive control' OR 'self-control' OR 'self control':ti,ab,kw OR ('self':ti,ab,kw AND 'regulation':ti,ab,kw) OR 'self-regulation':ti,ab,kw OR 'working memory':ti,ab,kw OR 'inhibition':ti,ab,kw OR ('inhibitory':ti,ab,kw AND 'control':ti,ab,kw) OR 'cognitive flexibility':ti,ab,kw) | 1226 |
| MELDINE Ovid | (cerebral palsy OR (perinatal AND stroke)) AND (relat* OR associat* OR correlat*) AND (executive function OR "executive control" OR "self-regulation" OR cognition/ OR cognition OR ("executive".tw. AND "control".tw.) OR self-control OR ("self".tw. AND "regulation".tw.) OR "self-regulation".tw. OR "working memory" OR inhibition OR ("inhibitory".tw. AND "control".tw.) OR "cognitive flexibility".tw.) | 495 |
| Cohrane Library | ("cerebral palsy" OR ("perinatal" AND "stroke")) AND (relat* OR associat* OR correlat*) AND ("executive function" OR "executive control" OR "self-regulation" OR "cognition" OR "executive control" OR "self-control" OR ("self" AND "regulation") OR "self-regulation" OR "working memory" OR "inhibition" OR ("inhibitory" AND "control") OR "cognitive flexibility") | 340 |
| CINAHL | ("cerebral palsy" OR (perinatal AND stroke)) AND (relat* OR associat* OR correlat*) AND ("executive function" OR "executive control" OR "self-regulation" OR cognition OR "self-control" OR ("self" AND regulation) OR "working memory" OR inhibition OR ("inhibitory" AND control) OR "cognitive flexibility") | 362 |
| Web of Science | ("cerebral palsy" OR (perinatal AND stroke)) AND (relat* OR associat* OR correlat*) AND ("executive function" OR "executive control" OR "self-regulation" OR cognition OR "self-control" OR ("self" AND regulation) OR "working memory" OR inhibition OR ("inhibitory" AND control) OR "cognitive flexibility") | 766 |
| ERIC IES | cerebral palsy AND executive functions OR executive function OR inhibition OR inhibitory control OR working memory OR cognitive flexibility OR self-regulation | 20 |
| APA PsycArticles | ("cerebral palsy" OR (perinatal AND stroke)) AND (relat* OR associat* OR correlat*) AND ("executive function" OR "executive control" OR "self-regulation" OR cognition OR "self-control" OR ("self" AND regulation) OR "working memory" OR inhibition OR ("inhibitory" AND control) OR "cognitive flexibility") | 558 |

Abbreviations: EMBASE, Excerpta Medica dataBASE; CINAHL, Cumulative Index to Nursing and Allied Health Literature; ERIC IES, Education Resources Information Center Institute of Education Sciences.

**Table S2.** Overview of tools assessing executive functions in the included studies

| Domain | Assessment tool | Type |
|---|---|---|
| Inhibitory control or Inhibition | Anti-saccade task | Performance-based test |
| | Day-Night task | Performance-based test |
| | Emotion regulation checklist | Questionnaire |
| | Stroop | Performance-based test |
| | Inhibit subscale, Behavior Rating Inventory of Executive Function | Questionnaire |
| | Inhibition index of Five Digit Test | Performance-based test |
| | Inhibition adapted by Jenks et al., 2009a | Performance-based test |
| | Knock-tap task | Performance-based test |
| | Inhibition, Naming condition, NEPSY | Performance-based test |
| | Inhibition, Inhibition condition, NEPSY | Performance-based test |
| | Auditory attention, NEPSY | Performance-based test |
| | Stop signal task | Performance-based test |
| Cognitive flexibility or Shifting | Animal Sorting, NEPSY | Performance-based test |
| | Shift subscale, Behavior Rating Inventory of Executive Function | Questionnaire |
| | Inhibition, Switching condition, NEPSY | Performance-based test |
| | Response set, NEPSY | Performance-based test |
| | Shifting adapted by Jenks et al., 2009a | Performance-based test |
| | Word generation task, NEPSY | Performance-based test |
| | Plus-Minus Task / Jersild Task | Performance-based test |
| | Trail-making Test | Performance-based test |
| | Wisconsin Card Sorting Test | Performance-based test |
| | Design Fluency | Performance-based test |
| | Lexical verbal fluency test | Performance-based test |
| Working memory or Updating | Corsi block-tapping test – backward condition | Performance-based test |
| | Backward Digit span, WISC | Performance-based test |
| | Backward Digit test, WMTB-C | Performance-based test |
| | Backward Spatial Span, WISC | Performance-based test |
| | Forced-choice recognition task | Performance-based test |
| | Infant working memory test | Performance-based test |
| | Letter-Number sequencing | Performance-based test |
| | Running Memory Paradigm / Morris & Jones Task | Performance-based test |
| | Working memory subscale, Behavior Rating Inventory of Executive Function | Questionnaire |
| Higher order executive functions (Reasoning, Planning, Problem-solving) | Balloon analogue risk task | Performance-based test |
| | Be a Communicator Construction | Performance-based test |
| | Be a Communicator Description without naming | Performance-based test |
| | Stockings of Cambridge | Performance-based test |
| | Tower of London, D-KEFS | Performance-based test |
| | Tower test, NEPSY | Performance-based test |
| | Rey/Osterrieth Complex Figure task | Performance-based test |
| | Planning and Organisation subscale, Behavior Rating Inventory of Executive Function | Questionnaire |
| Executive function composite | Global Executive Composite index, Behavior Rating Inventory of Executive Function | Questionnaire |
| | Five to Fifteen parental questionnaire | Questionnaire |
| | Executive function domain, Executive Function and Occupational Routines Scale | Questionnaire |
| | Composite score from multiple executive function tools | Performance-based test |

**Table S3.** Quality in Prognosis studies (QUIPS) checklist[1]

| | | | | |
|---|---|---|---|---|
| **Author and year of publication** | | | | |
| **Study identifier**<br>**Reviewer** | | | | |
| **Biases** | **Issues to consider for judging overall rating of "Risk of bias"** | **Study Methods & Comments** | **Rating of reporting** | **Rating of "Risk of bias"** |
| Instructions to assess the risk of each potential bias: | These issues will guide your thinking and judgment about the overall risk of bias within each of the 6 domains. Some 'issues' may not be relevant to the specific study or the review research question. These issues are taken together to inform the overall judgment of potential bias for each of the 6 domains. | Provide comments or text exerpts in the white boxes below, as necessary, to facilitate the consensus process that will follow. | Click on each of the blue cells and choose from the drop down menu to rate the adequacy of reporting as yes, partial, no or unsure. | Click on the green cells; choose from the drop-down menu to rate potential risk of bias for each of the 6 domains as High, Moderate, or Low considering all relevant issues |
| **1. Study Participation** | **Goal: To judge the risk of selection bias (likelihood that relationship between *PF* and *outcome* is different for participants and eligible non-participants).** | | | |
| *Source of target population* | The source population or population of interest is adequately described for key characteristics (LIST). | | | |
| *Method used to identify population* | The sampling frame and recruitment are adequately described, including methods to identify the sample sufficient to limit potential bias (number and type used, e.g., referral patterns in health care) | | | |
| *Recruitment period* | Period of recruitment is adequately described | | | |
| *Place of recruitment* | Place of recruitment (setting and geographic location) are adequately described | | | |
| *Inclusion and exclusion criteria* | Inclusion and exclusion criteria are adequately described (e.g., including explicit diagnostic criteria or "zero time" description). | | | |
| *Adequate study participation* | There is adequate participation in the study by eligible individuals | | | |
| *Baseline characteristics* | The baseline study sample (i.e., individuals entering the study) is adequately described for key characteristics (LIST). | | | |
| **Summary Study participation** | **The study sample represents the population of interest on key characteristics, sufficient to limit potential bias of the observed relationship between PF and outcome.** | | | |
| **2. Study Attrition** | **Goal: To judge the risk of attrition bias (likelihood that relationship between *PF* and *outcome* are different for completing and non-completing participants).** | | | |
| *Proportion of baseline sample available for analysis* | Response rate (i.e., proportion of study sample completing the study and providing outcome data) is adequate. | | | |
| *Attempts to collect information on participants who dropped out* | Attempts to collect information on participants who dropped out of the study are described. | | | |
| *Reasons and potential impact of subjects lost to follow-up* | Reasons for loss to follow-up are provided. | | | |
| *Outcome and prognostic factor information on those lost to follow-up* | Participants lost to follow-up are adequately described for key characteristics (LIST). | | | |
| | There are no important differences between key characteristics (LIST) and outcomes in participants who completed the study and those who did not. | | | |
| **Study Attrition Summary** | **Loss to follow-up (from baseline sample to study population analyzed) is not associated with key characteristics (i.e., the study data adequately represent the sample) sufficient to limit potential bias to the observed relationship between PF and outcome.** | | | |

| | |
|---|---|
| **3. Prognostic Factor Measurement** | **Goal: To judge the risk of measurement bias related to how PF was measured (differential measurement of PF related to the level of outcome).** |
| *Definition of the PF* | A clear definition or description of 'PF' is provided (e.g., including dose, level, duration of exposure, and clear specification of the method of measurement). |
| *Valid and Reliable Measurement of PF* | Method of PF measurement is adequately valid and reliable to limit misclassification bias (e.g., may include relevant outside sources of information on measurement properties, also characteristics, such as blind measurement and limited reliance on recall). |
| | Continuous variables are reported or appropriate cut-points (i.e., not data-dependent) are used. |
| *Method and Setting of PF Measurement* | The method and setting of measurement of PF is the same for all study participants. |
| *Proportion of data on PF available for analysis* | Adequate proportion of the study sample has complete data for PF variable. |
| *Method used for missing data* | Appropriate methods of imputation are used for missing 'PF' data. |
| **PF Measurement Summary** | ***PF* is adequately measured in study participants to sufficiently limit potential bias.** |
| **4. Outcome Measurement** | **Goal: To judge the risk of bias related to the measurement of outcome (differential measurement of outcome related to the baseline level of PF).** |
| *Definition of the Outcome* | A clear definition of outcome is provided, including duration of follow-up and level and extent of the outcome construct. |
| *Valid and Reliable Measurement of Outcome* | The method of outcome measurement used is adequately valid and reliable to limit misclassification bias (e.g., may include relevant outside sources of information on measurement properties, also characteristics, such as blind measurement and confirmation of outcome with valid and reliable test). |
| *Method and Setting of Outcome Measurement* | The method and setting of outcome measurement is the same for all study participants. |
| **Outcome Measurement Summary** | ***Outcome of interest* is adequately measured in study participants to sufficiently limit potential bias.** |
| **5. Study Confounding** | **Goal: To judge the risk of bias due to confounding (i.e. the effect of PF is distorted by another factor that is related to PF and outcome).** |
| *Important Confounders Measured* | All important confounders, including treatments (key variables in conceptual model: LIST), are measured. |
| *Definition of the confounding factor* | Clear definitions of the important confounders measured are provided (e.g., including dose, level, and duration of exposures). |
| *Valid and Reliable Measurement of Confounders* | Measurement of all important confounders is adequately valid and reliable (e.g., may include relevant outside sources of information on measurement properties, also characteristics, such as blind measurement and limited reliance on recall). |
| *Method and Setting of Confounding Measurement* | The method and setting of confounding measurement are the same for all study participants. |
| *Method used for missing data* | Appropriate methods are used if imputation is used for missing confounder data. |
| *Appropriate Accounting for Confounding* | Important potential confounders are accounted for in the study design (e.g., matching for key variables, stratification, or initial assembly of comparable groups). |
| | Important potential confounders are accounted for in the analysis (i.e., appropriate adjustment). |
| **Study Confounding Summary** | **Important potential confounders are appropriately accounted for, limiting potential bias with respect to the relationship between *PF* and *outcome*.** |
| **6. Statistical Analysis and Reporting** | **Goal: To judge the risk of bias related to the statistical analysis and presentation of results.** |
| *Presentation of analytical strategy* | There is sufficient presentation of data to assess the adequacy of the analysis. |
| *Model development strategy* | The strategy for model building (i.e., inclusion of variables in the statistical model) is appropriate and is based on a conceptual framework or model. |
| | The selected statistical model is adequate for the design of the study. |
| *Reporting of results* | There is no selective reporting of results. |
| **Statistical Analysis and Presentation Summary** | **The statistical analysis is appropriate for the design of the study, limiting potential for presentation of invalid or spurious results.** |

**Table S4.** Reason for studies' exclusion in the full-text screening phase

| | **Study (First author, year)** | **Reason for exclusion** | **Reference** |
|---|---|---|---|
| **1** | Ahlin et al. 2017 | No correlation coefficients between executive functions - ICF domains | [2] |
| **2** | Al-Nemr et al. 2018 | No correlation coefficients between executive functions - ICF domains | [3] |
| **3** | Anderson et al. 2009 | No correlation coefficients between executive functions - ICF domains | [4] |
| **4** | Araneda et al. 2021 | No correlation coefficients between executive functions - ICF domains | [5] |
| **5** | Bartonek et al. 2021 | No correlation coefficients between executive functions - ICF domains | [6] |
| **6** | Belmonti et al. 2015 | No correlation coefficients between executive functions - ICF domains | [7] |
| **7** | Belmonti et al. 2015a | No correlation coefficients between executive functions - ICF domains | [8] |
| **8** | Bosenbark et al. 2017 | Cerebral palsy population <80% | [9] |
| **9** | Busboom et al. 2024 | No correlation coefficients between executive functions - ICF domains | [10] |
| **10** | Coceski et al. 2021 | No correlation coefficients between executive functions - ICF domains | [11] |
| **11** | Crichton et al. 2020 | No correlation coefficients between executive functions - ICF domains | [12] |
| **12** | Di Lorenzo et al. 2022 | Cerebral palsy population <80% | [13] |
| **13** | Garcia-Castro et al. 2024 | No correlation coefficients between executive functions - ICF domains | [14] |
| **14** | Garcia-Galant et al. 2023 | No correlation coefficients between executive functions - ICF domains | [15] |
| **15** | Heyn et al. 2023 | No correlation coefficients between executive functions - ICF domains | [16] |
| **16** | Hoffman et al. 2021 | No correlation coefficients between executive functions - ICF domains | [17] |
| **17** | Kavcic et al. 2024 | Cerebral palsy population <80% | [18] |
| **18** | Koopmans et al. 2022 | No correlation coefficients between executive functions - ICF domains | [19] |
| **19** | Krivitzky et al. 2019 | Cerebral palsy population <80% | [20] |
| **20** | Laporta-Hoyos et al. 2022 | No correlation coefficients between executive functions - ICF domains | [21] |
| **21** | Mak et al. 2018 | No correlation coefficients between executive functions - ICF domains | [22] |
| **22** | Mak et al. 2022 | No correlation coefficients between executive functions - ICF domains | [23] |
| **23** | Micheletti et al. 2024 | No correlation coefficients between executive functions - ICF domains | [24] |
| **24** | Pagnozzi et al. 2016a | No correlation coefficients between executive functions - ICF domains | [25] |
| **25** | Pagnozzi et al. 2017 | No correlation coefficients between executive functions - ICF domains | [26] |
| **26** | Perro et al. 2006 | No correlation coefficients between executive functions - ICF domains | |
| **27** | Sakash et al. 2018 | No correlation coefficients between executive functions - ICF domains | [27] |
| **28** | Stadskleiv et al. 2016 | Domains of executive functions were correlated with other domains of EF | [28] |
| **29** | Tinderholt Myrhaug 2014 | No correlation coefficients between executive functions - ICF domains | [29] |
| **30** | van Abswoude et al. 2015 | No correlation coefficients between executive functions - ICF domains | [30] |
| **31** | Westmacott et al. 2009 | Inability to clarify study characteristics upon contact | [31] |
| **32** | White et al. 1994 | No correlation coefficients between executive functions - ICF domains | [32] |
| **33** | White et al. 2005 | No correlation coefficients between executive functions - ICF domains | [33] |
| **34** | Wilson et al. 2024 | Inability to clarify study characteristics upon contact | [34] |
| **35** | Wotherspoon et al. 2024 | No correlation coefficients between executive functions - ICF domains | [35] |
| **36** | Wotherspoon et al. 2024a | No correlation coefficients between executive functions - ICF domains | [36] |
| **37** | Yang et al. 2024 | Inability to obtain full record upon contact | |
| **38** | Zielinski et al. 2014 | No correlation coefficients between executive functions - ICF domains | [37] |

Abbreviations: ICF, International Classification System of Functioning and Disability; EF, executive functions.

**Table S5.** Overview of the 38 studies included in the systematic review and meta-analysis

| | First author. year | Title | Study design | Reference |
|---|---|---|---|---|
| **1** | Akyurek et al. 2024 | Comparison of the Executive Functions, Occupational Performance and Perceived Occupational Proficiency in Children with Neurodevelopmental Disorder | cross-sectional study | 38 |
| **2** | Ballester-Plane et al. 2018 | Cognitive functioning in dyskinetic cerebral palsy: Its relation to motor function, communication and epilepsy | case-control study | 39 |
| **3** | Belmonte Darraz et al. 2021 | Alteration of Emotion Knowledge and Its Relationship with Emotion Regulation and Psychopathological Behavior in Children with Cerebral Palsy | cross-sectional study | 40 |
| **4** | Cabezas & Carriedo 2020 | Inhibitory control and temporal perception in cerebral palsy | cross-sectional study | 41 |
| **5** | Caillies et al. 2012 | Theory of mind and irony comprehension in children with cerebral palsy | cross-sectional study | 42 |
| **6** | Critten et al. 2024 | Reading Disabilities in Children with Cerebral Palsy: associations with Working Memory | group case study | 43 |
| **7** | Dourado et al. 2013 | Association between executive/attentional functions and caries in children with cerebral palsy | cross-sectional study | 44 |
| **8** | Di Lieto et al. 2017 | Spastic diplegia in preterm-born children: Executive function impairment and neuroanatomical correlates | cross-sectional study | 45 |
| **9** | Forsman & Eliasson 2016 | Strengths and challenges faced by school-aged children with unilateral CP described by the Five To Fifteen parental questionnaire | cross-sectional study | 46 |
| **10** | Freire & Osoro 2023 | Executive functions and drawing in young children with cerebral palsy: Comparisons with typical development | cross-sectional study | 47 |
| **11** | Garcia-Galant et al. 2024 | Understanding social cognition in children with cerebral palsy: exploring the relationship with executive functions and the intervention outcomes in a randomized controlled trial | randomized controlled trial | 48 |
| **12** | Goble et al. 2012 | The influence of spatial working memory on ipsilateral remembered proprioceptive matching in adults with cerebral palsy | cross-sectional study | 49 |
| **13** | Jenks et al. 2007 | The Effect of Cerebral Palsy on Arithmetic Accuracy is Mediated by Working Memory, Intelligence, Early Numeracy, and Instruction Time | longitudinal study | 50 |
| **14** | Jenks et al. 2009 | Arithmetic difficulties in children with cerebral palsy are related to executive function and working memory | longitudinal study | 51 |
| **15** | Jenks et al. 2011 | Cognitive correlates of mathematical achievement in children with cerebral palsy and typically developing children | longitudinal study | 52 |
| **16** | Kalkantzi et al. 2025 | Daily-life executive functions and bimanual performance in children with unilateral cerebral palsy | cross-sectional study | 53 |
| **17** | Khan et al. 2021 | Executive behavior and functional abilities in children with perinatal stroke and the associated caregiver impact | cross-sectional study | 54 |
| **18** | Laporta-Hoyos et al. 2017 | White matter integrity in dyskinetic cerebral palsy: Relationship with intelligence quotient and executive function | cross-sectional study | 55 |
| **19** | Laporta-Hoyos et al. 2017a | Proxy-reported quality of life in adolescents and adults with dyskinetic cerebral palsy is associated with executive functions and cortical thickness | cross-sectional study | 56 |
| **20** | Laporta-Hoyos et al. 2018 | Brain lesion scores obtained using a simple semi-quantitative scale from MR imaging are associated with motor function, communication and cognition in dyskinetic cerebral palsy | cross-sectional study | 57 |
| **21** | Larsen et al. 2022 | Frontal interhemispheric structural connectivity, attention, and executive function in children with perinatal stroke | cross-sectional study | 58 |

| | | | | |
|---|---|---|---|---|
| **22** | Li et al. 2014 | The link between impaired theory of mind and executive function in children with cerebral palsy | cross-sectional study | 59 |
| **23** | Li et al. 2022 | Effects of Perinatal Stroke on Executive Functioning and Mathematics Performance in Children | cross-sectional study | 60 |
| **24** | Li et al. 2025 | Executive function is associated with behaviour problems in children and adolescents with cerebral palsy and intellectual disability | cross-sectional study | 61 |
| **25** | Meghji et al. 2024 | Executive functioning, ADHD symptoms and resting state functional connectivity in children with perinatal stroke | cross-sectional study | 62 |
| **26** | Mousavi et al. 2023 | A Study of the Relationship between Executive Function and School Function in Children with Cerebral Palsy | cross-sectional study | 63 |
| **27** | Nordberg et al. 2015 | Story retelling and language ability in school-aged children with cerebral palsy and speech impairment | cross-sectional study | 64 |
| **28** | Pagnozzi et al. 2016 | Automated, quantitative measures of grey and white matter lesion burden correlates with motor and cognitive function in children with unilateral cerebral palsy | cross-sectional study | 65 |
| **29** | Peeters et al. 2009 | Predictors of verbal working memory in children with cerebral palsy | cross-sectional study | 66 |
| **30** | Pirila et al. 2004 | A Retrospective Neurocognitive Study in Children With Spastic Diplegia | cross-sectional study | 67 |
| **31** | Soriano et al. 2021 | Speech-Language Profile Groups in School Aged Children with Cerebral Palsy: Nonverbal Cognition, Receptive Language, Speech Intelligibility, and Motor Function | cross-sectional study | 68 |
| **32** | Stadskleiv et al. 2014 | Investigating executive functions in children with severe speech and movement disorders using structured tasks | cross-sectional study | 69 |
| **33** | Stadskleiv et al. 2017 | Executive Functioning in Children Aged 6–18 Years with Cerebral Palsy | cross-sectional study | 70 |
| **34** | van Rooijen et al. 2012 | Arithmetic performance of children with cerebral palsy: The influence of cognitive and motor factors | cross-sectional study | 71 |
| **35** | van Rooijen et al. 2015 | From numeracy to arithmetic: Precursors of arithmetic performance in children with cerebral palsy from 6 till 8 years of age | longitudinal study | 72 |
| **36** | van Rooijen et al. 2016 | Working memory and fine motor skills predict early numeracy performance of children with cerebral palsy | cross-sectional study | 73 |
| **37** | Warschausky et al. 2017 | Mastery Motivation and Executive Functions as Predictors of Adaptive Behavior in Adolescents and Young Adults With Cerebral Palsy or Myelomeningocele | cross-sectional study | 74 |
| **38** | Whittingham et al. 2014 | Everyday psychological functioning in children with unilateral cerebral palsy: does executive functioning play a role? | cross-sectional study | 75 |

**Figure 1.** Funnel plots exhibiting publication bias

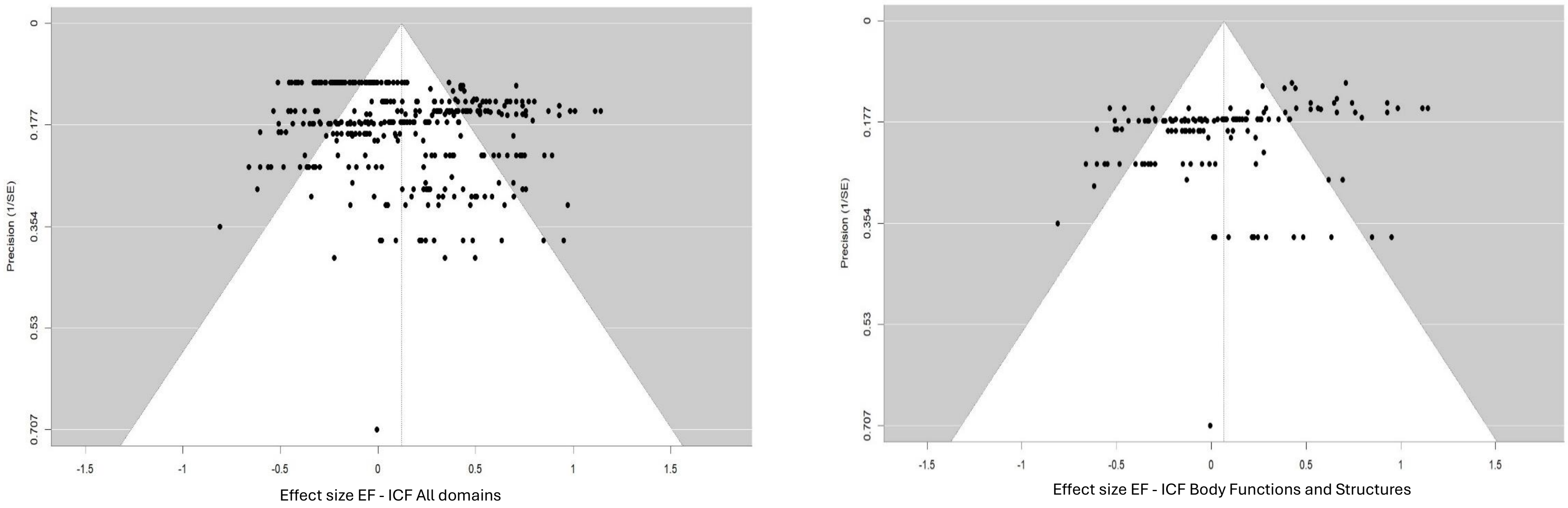

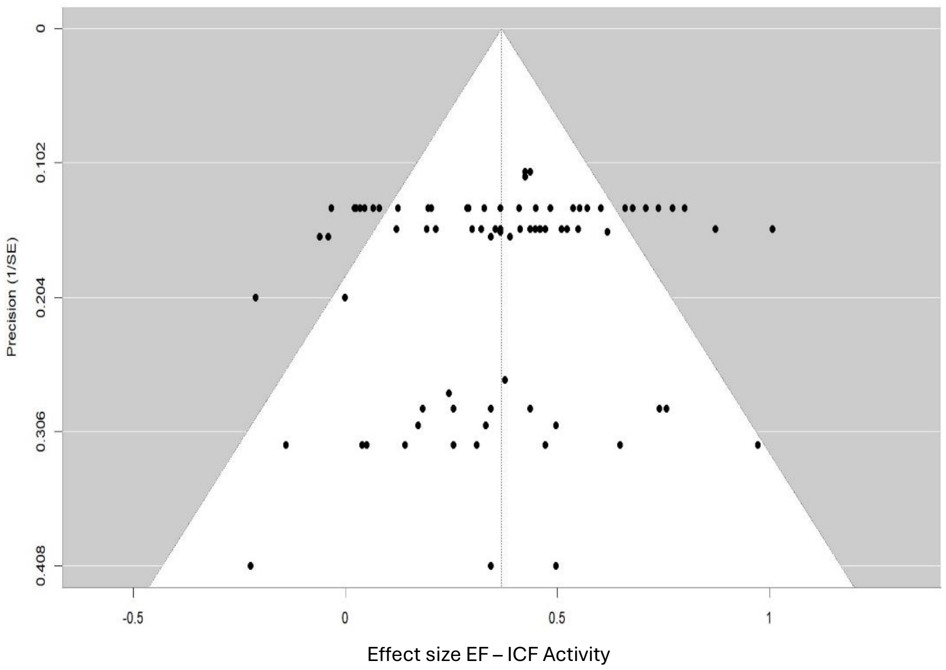
Precision (1/SE)
0
0.102
0.204
0.306
0.408
-0.5
0
0.5
1
Effect size EF – ICF Activity

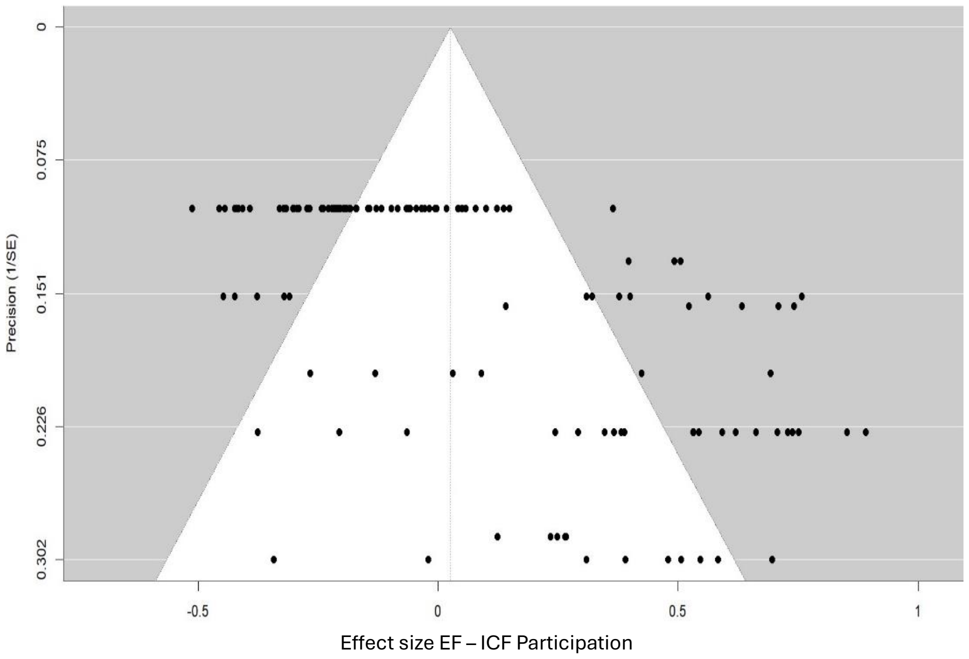
Precision (1/SE)
0
0.075
0.151
0.226
0.302
-0.5
0
0.5
1
Effect size EF – ICF Participation
